# Personalized virtual brains of Alzheimer’s Disease link dynamical biomarkers of fMRI with increased local excitability

**DOI:** 10.1101/2023.01.11.23284438

**Authors:** Bahar Hazal Yalçınkaya, Abolfazl Ziaeemehr, Jan Fousek, Meysam Hashemi, Mario Lavanga, Ana Solodkin, Anthony R. McIntosh, Viktor K. Jirsa, Spase Petkoski

## Abstract

Alzheimer’s disease (AD) is a neurodegenerative disorder characterized by the accumulation of abnormal beta-amyloid (Aβ) and hyperphosphorylated Tau (pTau). These proteinopathies disrupt neuronal activity, causing, among others, an excessive and hypersynchronous neuronal firing that promotes hyperexcitability and leads to brain network dysfunction and cognitive deficits. In this study, we used computational network modeling to build a causal inference framework to explain AD-related abnormal brain activity. We constructed personalized brain network models with a set of working points to enable maximum dynamical complexity for each brain. Structural brain topographies were combined, either with excitotoxicity, or postsynaptic depression, as two leading mechanisms of the Aβ and pTau on neuronal activity. By applying various levels of these putative mechanisms to the limbic regions that typically present, with the earliest and largest protein burden, we found that the excitotoxicity is sufficient and necessary to reproduce empirical biomarkers two biometrics associated with AD pathology: homotopic dysconnectivity and a decrease in limbic network dynamical fluidity. This observation was shown not only in the clinical groups (aMCI and AD), but also in healthy subjects that were virtually-diseased with excitotoxicity as these abnormal proteins can accumulate before the appearance of any cognitive changes. The same findings were independently confirmed by a mechanistic deep learning inference framework. Taken together, our results show the crucial role of protein burden-induced hyperexcitability in altering macroscopic brain network dynamics, and offer a mechanistic link between structural and functional biomarkers of cognitive dysfunction due to AD.

## Introduction

Alzheimer’s disease (AD) is a neurodegenerative disorder characterized by the progressive loss of cognitive function, particularly memory and other cognitive abilities. It is marked by the accumulation of extracellular beta-amyloid plaques (Aβ) and intracellular hyperphosphorylated tau-forming neurofibrillary tangles (NFT), which leads to the progressive neuronal degeneration of the cortical and subcortical structures. The spatial and temporal distribution of these neuropathological changes follow a pattern originally described by Braak and Braak system (Braak & Braak, 1991) (Figure 1A), where the accumulation of Aβ concentrate first appear in temporal and frontal regions before spreading out to other association areas followed by primary areas along the ventromedial regions and finally basal ganglia. In contrast, pTau first accumulates in ventromedial temporal areas followed by other temporal regions before spreading into association cortices before reaching the primary cortical regions (Figure 1B).

**Figure 1.**
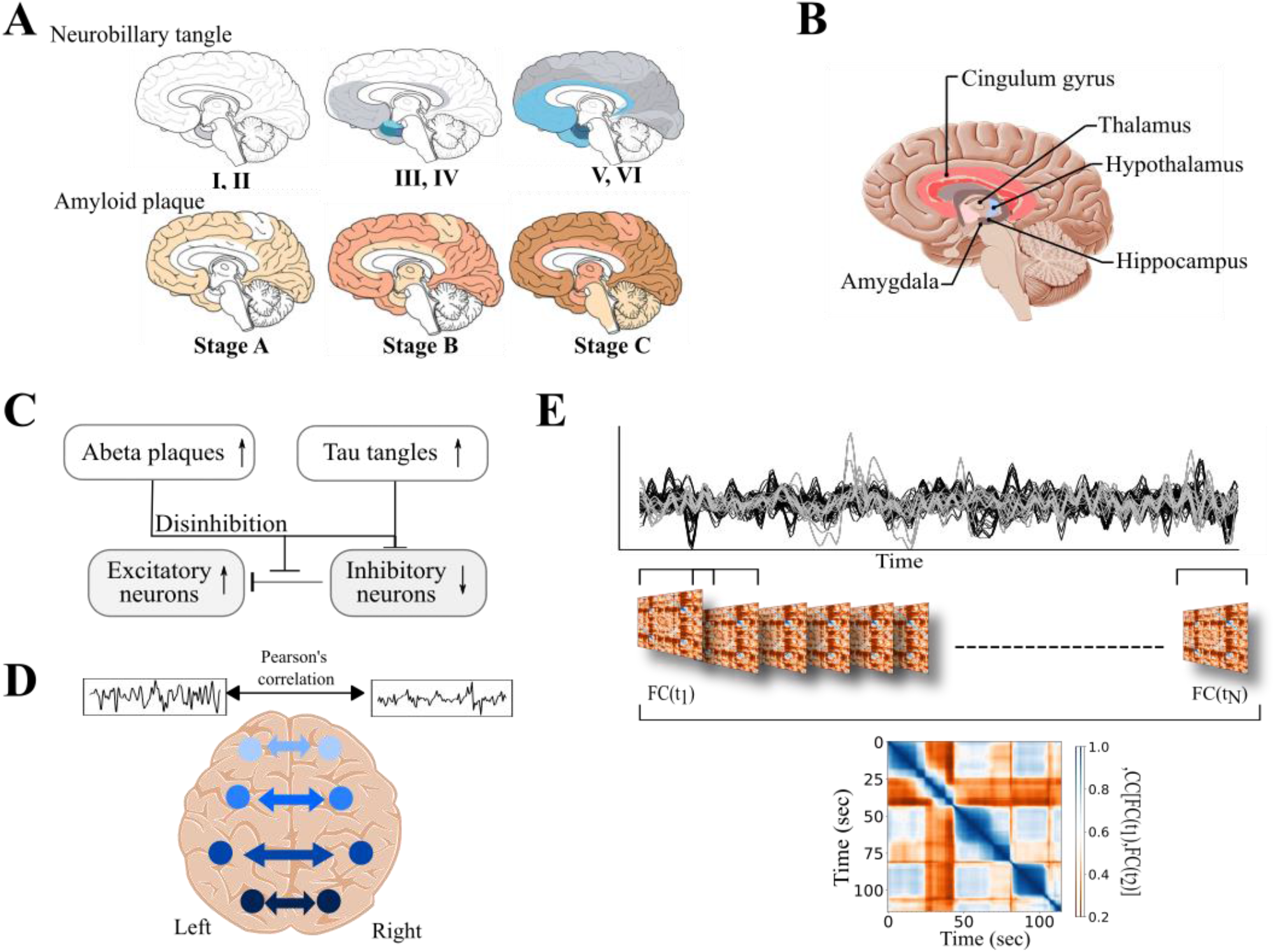
Schematic description of reliable summary statistics in AD.**A**. Spatiotemporal evolution of Aβ and pTau deposits in AD (Braak and Braak, 1991). **B**. The regions of the limbic subnetwork. **C**. Neuropathological alterations of Aβ and pTau disrupts synaptic transmission, perturbs excitation/inhibition balance and alters neuronal networks resulting in the excitotoxicity that leads cognitive decline in AD. **D**. Representative image for Homotopic FC: mirror connections between right and left hemisphere. **E**. Static and dynamic functional connectivity matrices derived from rs-fMRI time series. Static FC was calculated using Pearson’s correlation coefficients of the entire time series, whereas FCD was calculated by sliding a temporal window over the time series.

The characteristic accumulation of both Aβ and pTau are considered diagnostic neuropathological markers for AD. Interestingly, the progressive accumulation of pTau pathology in plaque-bearing areas occurs around Braak’s stage III and has been suggested as the to a shift from the asymptomatic preclinical stage to the symptomatic clinical stage of the disease (Delacourte et al,. 1991, Musiek and Holtzman., 2015, Wang et al,. 2016, Pontecorvo et al,. 2017, Ossenkoppele et al., 2022). The extent of AD-related pathology in the brain can be assessed using positron emission tomography (PET) scans and cerebrospinal fluid (CSF) assays, along degeneration at the structural level and functional changes detected with functional magnetic resonance imaging (fMRI) and non-invasive electromagnetic recording techniques such as electroencephalography (EEG) and magnetoencephalography (MEG).

The neuropathological changes in brain due to proteinopathies, has been suggested can lead to impaired communication between neurons and a consequent dysfunction in neural networks, resulting in cognitive deficits (DeTrue and Dickson, 2019). Early research in AD demonstrated the association between NFT with the progressive loss of neurons and impaired synapses, leading to overt atrophy in cortical and subcortical areas (Delbeuck et al,. 2003, Catani and Ffytche, 2005).

However, more recent studies have shown that before clear degeneration is observed, there are more subtle functional changes associated with detected as an imbalance in excitation and inhibition (E/I) in the brain possibly due to the impairment of inhibitory neurons (Schmid et al., 2016, Busche and Konnerth, 2016; Ambrad Giovannetti and Fuhrmann, 2019; Hijazi et al., 2019), loss of inhibitory receptors and synapses, and disruption of glutamate reuptake (Zott et al., 2019). Interestingly, interneurons do not develop NFTs and seem to degenerate trans-synaptically as a consequence of degeneration of principal neurons (Solodkin et al., 1996). Initially, it was assumed that Aβ was solely responsible for the increase in neuronal excitability as it contributes to memory loss and degenerative changes. Now, AD is acknowledged as a tauopathy, and it has been established that it plays a major role in the hyperexcitability of certain brain regions (Roberson et al., 2007; Pooler et al., 2013; Wu et al., 2016). Furthermore, there have been findings that tau also causes hypoexcitability in certain areas of the brain (Busche et al., 2012; Hatch et al., 2017). Additionally, some studies have suggested that there is a synergistic relationship between Aβ and tau, with tau mediating the hyperexcitability caused by Aβ (Roberson et al., 2007; Ittner et al., 2010; DeVos et al., 2013). Nevertheless, the literature on the effects of AD proteinopathies on various cell types is contradictory, making it hard to comprehend the dynamics of the illness.

Neural hyperexcitability is an early symptom of the disease, and this makes excitotoxicity an attractive option for both therapeutic and modeling purposes. To address this, computational modeling can be used as a tool to investigate the link between microscale cellular mechanisms and macroscale abnormal neuronal dynamics in AD. Computational modeling allows us to simulate real-world systems, thus learning about the data generation process, making predictions on unseen data, and justifying the hypotheses. By running various simulations with parameters informed by the individual structural data and hypothesized mechanisms, we can gain insight into different potential outcomes, thus providing evidence to support or refute hypotheses regarding the effects and mechanisms of a certain phenomenon.

Modeling approaches that examine the whole-brain (Ghosh et al., 2008; Deco et al., 2011; Sanz Leon et al., 2013; Breakspear, 2017; Bassett et al., 2018; Lavanga et al., 2021) can characterize the relationship between brain anatomy and brain dynamics, allowing for the prediction of the effects based on the known causes. These models can also contain regionally variant parameters, when this is supported by clinical (Jirsa et al., 2017; Courtiol et al., 2020) or data informed (Kringelbach et al., 2020) hypotheses. In the context of AD, studies shifted their focus to linking local and global dynamics to the main driver of the disease to pathophysiology driven neuronal hyperactivity via The Virtual Brain platform (Sanz Leon et al., 2013; Zimmermann et al., 2018; Stefanovski et al., 2021; Triebkorn et al., 2022; Arbabyazd et al., 2021; Patow et al., 2022). These studies have demonstrated the value of this approach. Especially important results indicated in Triebkorn et al., 2022, showed the added values of the model for classification based on the structural and functional data features.

In contrast to forward modeling, which is a top-down approach, model inversion is a bottom-up strategy that infers hidden causes from observed effects (D’Angelo and Jirsa, 2022). However, performing whole-brain model inversion, which also allows for uncertainty quantification and the incorporation of prior information in hypothesis testing, is challenging (Hashemi et al., 2020) mainly due to the nonlinear and high-dimensional nature of latent dynamics (not directly measurable). This requires the integration of various sources of information, such as neural mass models, anatomical data, empirical observations, and advanced probabilistic machine learning techniques, in a unified framework called Bayesian inference (Friston 2010, Hashemi et al., 2020), as performed in this study. This approach allows for the incorporation of uncertainty and prior knowledge in the hypothesis testing process, which is important for understanding the underlying causes of AD.

Structural MRI and resting-state fMRI connectivity can serve as biomarker surrogates, providing insight into the characteristics of brain architecture and neural processes in AD (Yu et al., 2021). Structural connectivity (SC) is generally assessed using diffusion tensor imaging and can be represented as a connectivity matrix that characterizes the anatomical connections between brain regions through white matter tracts. Abnormalities in SC in AD patients are typically reported as changes in parameters of water diffusion, such as a significant decrease in fractional anisotropy and an increase in mean diffusivity in the whole-brain (Pievani et al., 2010, Huang et al., 2012, Acosta-Cabronero et al., 2012, Douaud, et al., 2013). In AD the severity of these measures of overall diffusivity in the tissue and the degree of directional restriction of water diffusion are correlated with the clinical phenotype and disease progression. Functional connectivity (FC) refers to the synchronization of neuronal activity across brain regions and is often measured using statistical techniques such as correlation. Resting-state FC analysis has shown alterations in AD, including decreased FC strength predominantly in areas with high protein burden and increased neural activity and loss of functional specificity in posterior-medial regions in individuals with cognitive impairment and mixed Aβ-pTau status (Maass et al., 2019). Specifically, reduced inter-hemispheric homotopic FC (Figure 1D) is also associated with AD (Wang et al., 2015, Qiu et al., 2016). Functional connectivity dynamics (FCD) (see Figure 1.E) refers to the flexibility of the brain to flow between different states to facilitate cognition and has been shown to be altered in healthy lifespan (Battaglia et al., 2020, Petkoski et al., 2022), normal mild-life and older adults (Lavanga et al., 2022), and AD (Córdova-Palomera et al., 2017, Jie et al., 2018). These changes in FCD may provide insight into the neural mechanisms underlying cognitive decline in AD.

Here we provide computational network modeling as a tool to establish a causal inference framework that could shed light on the abnormal brain activity seen in individuals suffering from AD. Diffusion-weighted imaging (DWI) data of 115 subjects (71 Healthy, 32 aMCI and 12 AD) and resting-state functional magnetic resonance imaging (rs-fMRI) data were investigated as two groups that are clinically categorized as healthy controls and cognitively impaired subjects (aMCI and AD patients). Specifically, in empirical recordings we observed changes in homotopic inter-hemispheric FC and FCD of the limbic subnetwork. In order to explain the empirical abnormal brain activity described above, our personalized brain network model incorporated structural brain topographies along with either excitotoxicity or postsynaptic depression. These two mechanisms are thought to be among the most influential ways in which proteinopathy affects neuronal activity. By applying two contrasting disease mechanisms: excitotoxicity and postsynaptic depression at different levels, we are able to investigate their direct affect on brain function. This approach allowed us to gain a better understanding of the pathological trajectory of AD. Through this process, we found that only excitotoxicity was sufficient and necessary to reproduce the empirical biomarkers of AD pathology. This finding held true not only in the diseased group, but also in healthy subjects who were virtually induced with excitotoxicity, revealing a possible disease trajectory in biophysically relevant parameters. These results suggest that excitotoxicity plays a crucial role in the development of AD pathology and may be an important target for therapeutic intervention. In addition, our findings confirm the importance of the limbic regions in the development of AD pathology and the potential impact of protein burden on brain function. In line with the recent evidence that cement the importance of Aβ and pTau in the limbic regions as AD biomarkers (Ossenkoppele et al., 2022), our approach provides a mechanistic link to the observed dynamical alterations. Additionally, as a validation step we adapt a mechanistic causal inference framework for the accurate estimation of pathological trajectory by training the deep neural density estimators on model simulations, and the low-dimensional empirical evidence. This is performed by embedding the state-of-the-art deep neural density estimators in simulation-based inference methods (Cranmer et al., 2022) to learn an invertible transformation between parameters and data features from a budget of simulations (Gonçalves 2020, Hashemi et al., 2022).

## Results

In this study, we explored structural and functional biomarkers that are available in AD. We built a mechanistic framework using TVB to create virtual brains with individual structural connectomes and used them to generate functional data ensembles to better understand the evolution of cognitive impairment in patients with aMCI and AD patients. Before introducing new features to models we investigated brain network models simulated data, which was constrained by only individual connectome. As expected, these were not sufficient to capture the representative dynamics of each group. To entangle the impact of the proteinopathies, Aβ and pTau, we conceptualized distinct hypotheses based on alterations shown in the limbic subnetwork: attenuated postsynaptic connections, and hyperexcitability. Simulations of personalized brains with hyperexcitability revealed that only the latter virtual cohort can describe the functional reorganization of oscillatory dynamics as observed in the static and dynamic FC metrics (Figures 2 B, C). We have verified that the white matter atrophy, as evidenced by empirical connectome, was not sufficient enough to be the sole casual mechanism of the functional alterations observed in the patients’ brains. Lastly, we independently validate the causal hypothesis of TVB with a pathology pipeline using the unified Bayesian inference framework.

**Figure 2.**
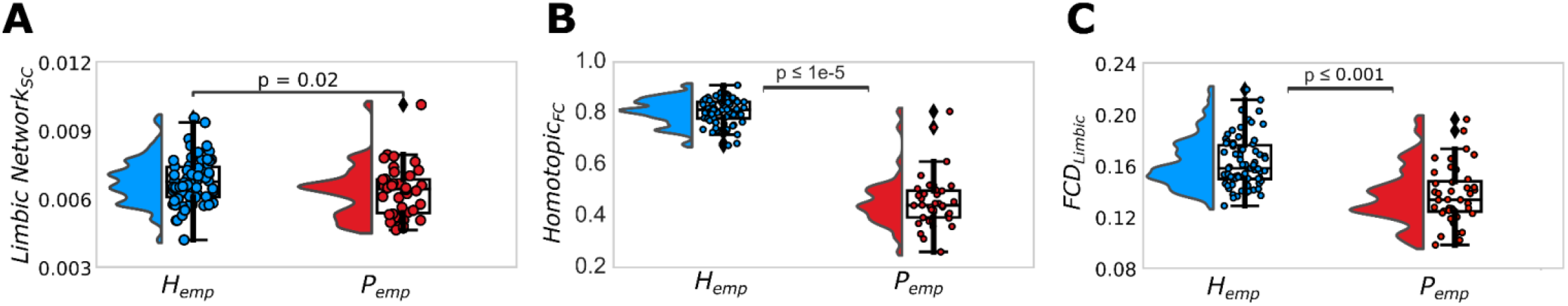
Significant structural and functional connectivity alterations in healthy and patient cohorts. (*P*_*emp*_: the patient group, *H*_*emp*_: the healthy group) **A**. Within limbic network averaged connections (*SC*_*emp*_) show the loss of links in cognitively declined individuals (*p* = *0*.*02*). **B**. Patient-specific functional organizations show a significant homotopic dysconnectivity (*p* ≤ *1e*^−*5*^). **C**. Brain’s FCD variances (fluidity) show significant reduction within the limbic regions in cognitively declined group compared to the healthy group (*p* ≤ *0*.*001*).

### R.1. Structural and Functional Connectivity analysis

Subjects’ structural phenotypes (*SC*_*emp*_) were derived from DWI data of the participants of the MAS study dataset (n=115, age-range between 70-90 years), using AAL atlas (88 regions) (for details see Material and Methods section). Both structural and functional changes have been observed in the brains of individuals with AD. Structural changes in AD include atrophy, or shrinkage, of brain regions such as the hippocampus (Josephs et al., 2017), a region important for memory consolidation, and the prefrontal cortex, a region involved in executive function (Poo et al., 2016). White matter atrophy is one of the common features of AD, which can disrupt the communication between different brain regions and contribute to the cognitive decline seen in this disorder. Studies have demonstrated that white matter atrophy is present in the brains of individuals with AD, both in early and advanced stages of the disease (Fjell et al., 2009; Jones et al., 2012). The extent of white matter atrophy has also been found to be correlated with the severity of cognitive impairment in AD (Fjell et al., 2009). Structural organization matrices (*SC*_*emp*_) were investigated in two clinical groups (Healthy control and Cognitively Declined representing aMCI and AD patients). The integrity testing of white matter connections across the groups showed system specific disease-related structural atrophy between the links of the region’s limbic network (Figure 2A, ANOVA: *F* = 5.55, *p* = 0.02), which was also reported in earlier work provided by Zimmerman et al, 2018. Connectivity of the other lobes did not show statistical difference between the groups (see Figure S1, and neither did the whole-brain, intra and inter-hemispheric structural connectivities, Figure S2).

AD is generally characterized by the progressive loss of cognitive function where the coordination of resting-state brain activity between brain regions is disrupted, particularly networks involved in emotion and memory processing. The summary statistics of the empirical resting-state fMRI to reveal functional criterions of pathology identified two data features: (i) functional homotopic dysconnectivity and (ii) dynamical variability. Homotopic functional connectivity is the key characteristic feature of intrinsic connection in the human brain, as its alteration has been reported in MCI and AD patients (Wang et al., 2015). We investigated homotopic FC to reveal the influence of pathogenesis of disease on co-activation between hemispheres. The homotopic FC is computed as the average of the Pearson correlation coefficient between the regions in one hemisphere and their mirrored regions in the other (e.g., prefrontal right and left), which obtained as the diagonal of order, with N as the total number of brain regions. In addition to abnormal FC, the changes in FCD are also reported in AD using different approaches (Jones et al. 2012; Cordova-Palomera et al. 2017; Demirtas et al. 2017). The temporal evolution of the brain FC is represented by sliding window technique and it is quantified via switching index (for further detail Material and Methods section). The index related with brain fluidity (Figure 1E) (Hansen et al., 2015; Melozzi et al., 2019; Courtiol et al., 2020) quantifies the switching behaviour of resting state networks (Rabuffo et al., 2021; Lavanga et al., 2022). Empirical resting-state fMRI analysis showed that patients with cognitive decline had a significant reduction in both homotopic dysconnectivity (Figure 2B), and brain fluidity as measured by FCD variance of the limbic network (Figure 2C) (Pillai’s Trace = 0.86, *F*(4, 218) = 40.97, *p* < 0.001). Supplementary Figure S1, B summarizes static within and between network FC results, which showed generally reduced connectivity within network, and increased between them.

### R.2 Virtual aging with pathology: the causal link between excitability and functional connectivity

Hyperexcitability, or excessive activity in the brain, has been suggested as a key pathogenesis that occurs in the brains of individuals with AD due to protein burden. This hyperexcitability is thought to contribute to the overall brain dysfunctions that are characteristic of AD including cognitive impairments, neurodegeneration, and inflammation. To investigate the potential impact of structural or molecular changes on brain function in AD, we created personalized virtual brains for each subject. We aimed to create an avatar brain that resembles the changes in brain dynamics in AD that originated because of structural or molecular changes. To generate simulated data, we used the open-source platform TVB to implement brain network models for each subject. This framework allows us to simulate patients and healthy connectomes by varying different hyperparameters such as, global modulation *G* that represents the impact of structural data over the whole-brain’s dynamics, and the noise variance *σ*^*2*^ that represents the stochastic fluctuations driven by a generic environmental noise to capture switching behavior of functional brain networks that reflects realistic evaluation of brain function (details can be found Rabuffo et al., 2021). The temporal evolution of simulated brain connectivity in connectome-based simulations was evaluated using the most realistic switching index couple (*G, σ*^*2*^). (The global modulation associated with maximum FCD variance of each subject and whole-brain FCD variance in the healthy controls compared to the cognitively declined group with pathology shown in Supplementary Figures S3 B,C). BNM simulated data with individual connectomes was not able to generate an ensemble of key features of empirical functional data (Figures 3A, B).

**Figure 3.**
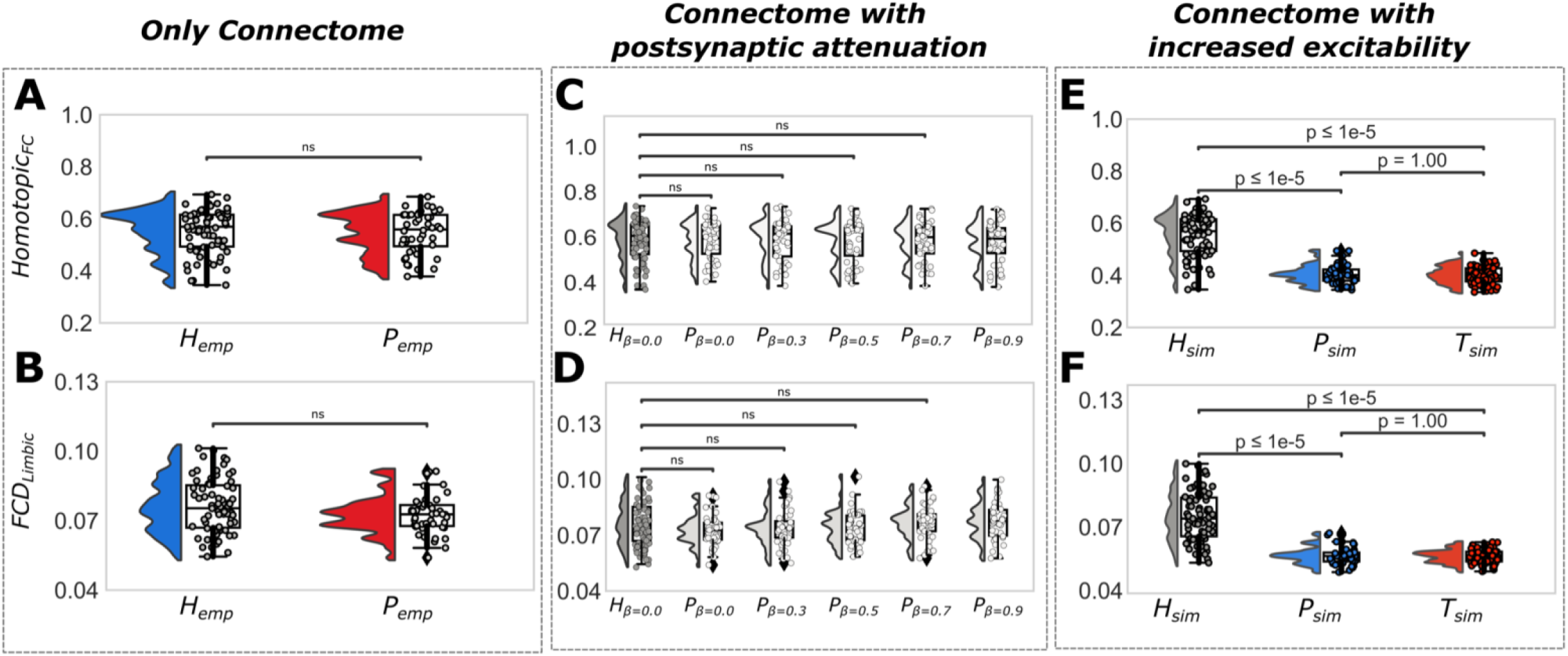
Testing the hypothesized mechanisms of BNMs on the homotopic FC and its dynamics.**A-B**. Simulated connectome-based model with subject-specific structural phenotype (*SC*_*emp*_) with no explicit impact of protein burden (neither excitotoxicity, or with postsynaptic depression) is not able to replicate the features that are significantly different in empirical clinical groups. **C-D**. Postsynaptic attenuation is achieved by artificial masking of the within limbic network and postsynaptic connections in the SC matrix (from 0% to 100% decrease). However, the homotopic functional dysconnectivity and decreased fluidity of the limbic subnetwork, that are seen in empirical clinical groups, were not replicated by the simulated data. **E-F**. Hyperexcitability is introduced via local excitability parameter into limbic subnetwork (the image represents when *η* = −*3*.*58* in the limbic subnetwork, where the default for other regions is *η* = −*4*.*7*). This is done for patients group (*P*_*sim*_) and for the virtually diseased group consisting of the healthy subjects with hypothesized impact of the protein burden (*T*_*sim*_). The local excitability parameter changed only in the regions within the limbic network to mimic the effect of hyperexcitability, specifically for these regions.

For causality testing, we hypothesized a distinct in-silico experiment that distinguishes the disease trajectory from normal aged brain dynamics. As suggested by previous works (Roberson et al., 2007, Ittner et al., 2010, DeVos et al., 2013), disease related excitotoxicity was shown in the protein accumulated brain areas. We casually test whether excitotoxicity of the limbic network regions over disease could be the main driving force of the empirically recorded dynamical activity difference between healthy controls and cognitively declined. To simulate the whole-brain dynamics we used BNMs introduced by Rabuffo et al., 2021, consisting of interconnected nodes through the SC matrix, each of which represents the average neural activity of infinite and all-to-all coupled quadratic integrate-and-fire neurons (Montbrió et al., 2015). The emergent brain dynamics are constrained by structural data (anatomical pathways), allowing the hidden brain states and the trajectories in the latent space to be inferred from the data, due to the coupling between brain regions.

The neural activity of the regional neuronal population is governed by the excitatory network with heterogeneous input 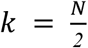, whereas the global modulation *G*. In BNM fitted by parameters *G* modulation index and excitatory limbic network with heterogeneous input 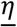 to capture switching behavior of functional brain networks that reflects realistic evaluation of brain function. After having the most realistic switching index couple 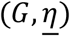 which is the point where each decoupled node in its bistable regime, in the 2D phase space demonstrates a ‘down’ fixed point and an ‘up’ stable focus (Rabuffo et al., 2021). This realistically represents the switching behavior observed in empirical data (Hansen et al., 2015; Rabuffo et al., 2021) and provides working points at the maximum fluidity.

All BNMs were simulated using the TVB platform, generating neural activity at each region of the whole-brain, mapped to the BOLD time series. The associated BOLD activity for each brain region obtained by filtering raw high time-resolution neuroelectric signals, through the Balloon-Windkessel model (Friston et al., 2000). We explore the parameter space, varying the heterogeneous input 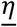 (excitability parameter of limbic network), the global parameters *G*, and the noise variance *σ*^*2*^, to best match the simulated BNM with empirical data.

Specifically, in-silico experiments were conducted in two scenarios: (i) to capture the effect of changes in heterogeneous input 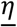 in the limbic network on whole-brain dynamics, while we constrained the brain network dynamics by the connectomes of the aMCI and Alzheimer’s subjects (labeled as *P*_*sim*_). We effectively explored the effects of increased excitability limbic network regions (−4.7 to -3.5) on functional data of these patients’ connectomes. (ii) to create our virtual diseased cohort, we constrained brain network models relying on the connectomes of the normal subjects (labeled as *T*_*sim*_), while increasing heterogeneous input 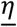 of the limbic network as the same level as it was applied in the first scenario. We explored the parameter space, varying the global parameter *G* and heterogeneous input of firing-rate equations 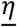. Each brain network model were tuned by sweeping the parameters in target range (*G* ∈ [*0*.*2, 1*.*1*], *η* ∈ [−*6*.*0*, −*3*.*5*], with *σ*^*2*^ = *0*.*04*) and those values are selected that maximize the variance of whole-brain FCD, *σ*_*full*_ ^*2*^ (see Methods). To verify that our virtual BNMs can generate disease related empirical functional changes, we monitored a set of biomarkers that represents the disease causing functional features. In terms of homotopic FC, the brain network model constrained by connectomes of the aMCI and Alzheimer’s patients, *P*_*sim*_, and the virtualized brain simulations, *T*_*sim*_, showed reduction with the increasing heterogeneous input 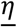(Figure 3E) compared to healthy simulated data. (Kruskal-Wallis paired samples with Bonferroni correction homotopic FC: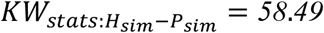 with 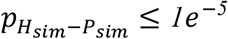, and 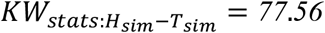 with 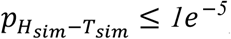. Similar to the homotopic FC, we observed an pathology related decline in the limbic subnetwork FCD variance difference (*σ*^*2*^_*FCD* −*limbic*_) in all two datasets (Figure 3F). (Kruskal-Wallis paired samples with Bonferroni correction limbic network FCD: 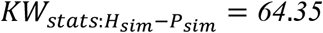 with 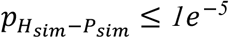, and 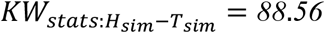 with 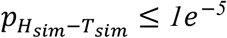. The effect of increasing heterogeneous input *η* of aMCI and Alzheimer’s patients brain dynamics compared to healthy control with *η* = −*4*.*7* in terms of the homotopic FC and limbic network fluidity shown in Supplementary Figure S4. Additionally, global modulation and fluidity whole brain dynamics measures in patients group (*P*_*sim*_) and for the virtually diseased group consisting of the healthy subjects with hypothesized impact of the protein burden (*T*_*sim*_) indicated when *η* = −*3*.*58* in Supplementary Figures S5 A,B, respectively.

As a validation to our virtual brain BNMs, we focused on the relation between anatomical and FC changes. We constrained the brain network model via the connectomes of the patients (connectomes of the aMCI and Alzheimer’s subjects), whose limbic network and postsynaptic connections SC were homogeneously decreased from 0% to 100% (details can be found in **SM-** Connectome-based model with structural impact of protein burden on limbic network and postsynaptic links, Figure S1). The constrained brain network model via decreasing connections limbic network and postsynaptic links failed to build a virtually aged brain network model with pathology (Figures 3C, D).

### R.3. Bayesian approach

Simulation-based inference (SBI) provides flexible and efficient Bayesian model inversion using only simulated samples from a complex probabilistic model, with low computational cost when standard methodologies cannot be applied (Cranmer et al., 2020). In this study, we used the sequential neural posterior estimation (SNPE; Gonçalves et al., 2020) algorithm from SBI methods to perform Bayesian model inversion, which provides us with an efficient direct estimation of the posterior distributions of BNM parameters, such as the global modulation *G* and excitability of limbic network regions 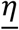. This results from updating the prior probability of having a plausible range over parameters with the information provided by empirical data, through the likelihood function, which in our case relies on personalized and mechanistic BNMs. To this end, we trained the masked autoregressive flows (MAFs; Papamakarios et al., 2017) as the state-of-the-art neural density estimator, on the data features of an ensemble of 65k model simulations, with random parameters. MAF is a class of deep neural networks that enables highly expressive transformations to quickly reconstruct complex probability density functions with low-cost computation. The simulation were performed with uniform prior as *G* ∈ [*0*.*2, 1*.*1*] for the global coupling parameter, and *η* ∈ [−*6*.*0*, −*3*.*5*] for the excitability parameter in limbic network regions. The data features include the homotopic dysconnectivity and brain FCD variance reduction in the limbic subnetwork (for further details see Material and Methods section). The uncertainty quantification shows that FCD variance is more informative than FC homotopic, but using both data features provides stronger model evidence against the fitted data (Figure S9).

For the validation of SBI approach, a set of synthetic data with known parameters was generated as the observations, and our results indicate that SBI is able to accurately estimate the joint posterior distribution of the parameters (*G, η*) that narrows around the true parameters (Supplementary Figure S7).

Using SBI on the cohort’s empirical data, the uncertainty in virtual brain trajectories was quantified through Bayesian estimation of parameters (*G, η*), for both healthy and AD groups. Figure 4 shows the pooled estimated posterior distributions of parameters *G* and *η* for each group, indicating non-significant change of the global modulation *G* over disease (*p* = *0*.*05*, Supplementary Figure S6). This was expected since no structural alterations were observed in patients’ connectomes rather than a decrease in the limbic network connections (Figure 2A). In contrast, the excitability parameter of limbic network regions is significantly increased for the patient cohort compared to the healthy controls (Figure 4A). This demonstrates that the change in excitability in those regions prone to protein accumulation is a relevant data feature that conveys the disease related empirically recorded functional changes.

**Figure 4.**
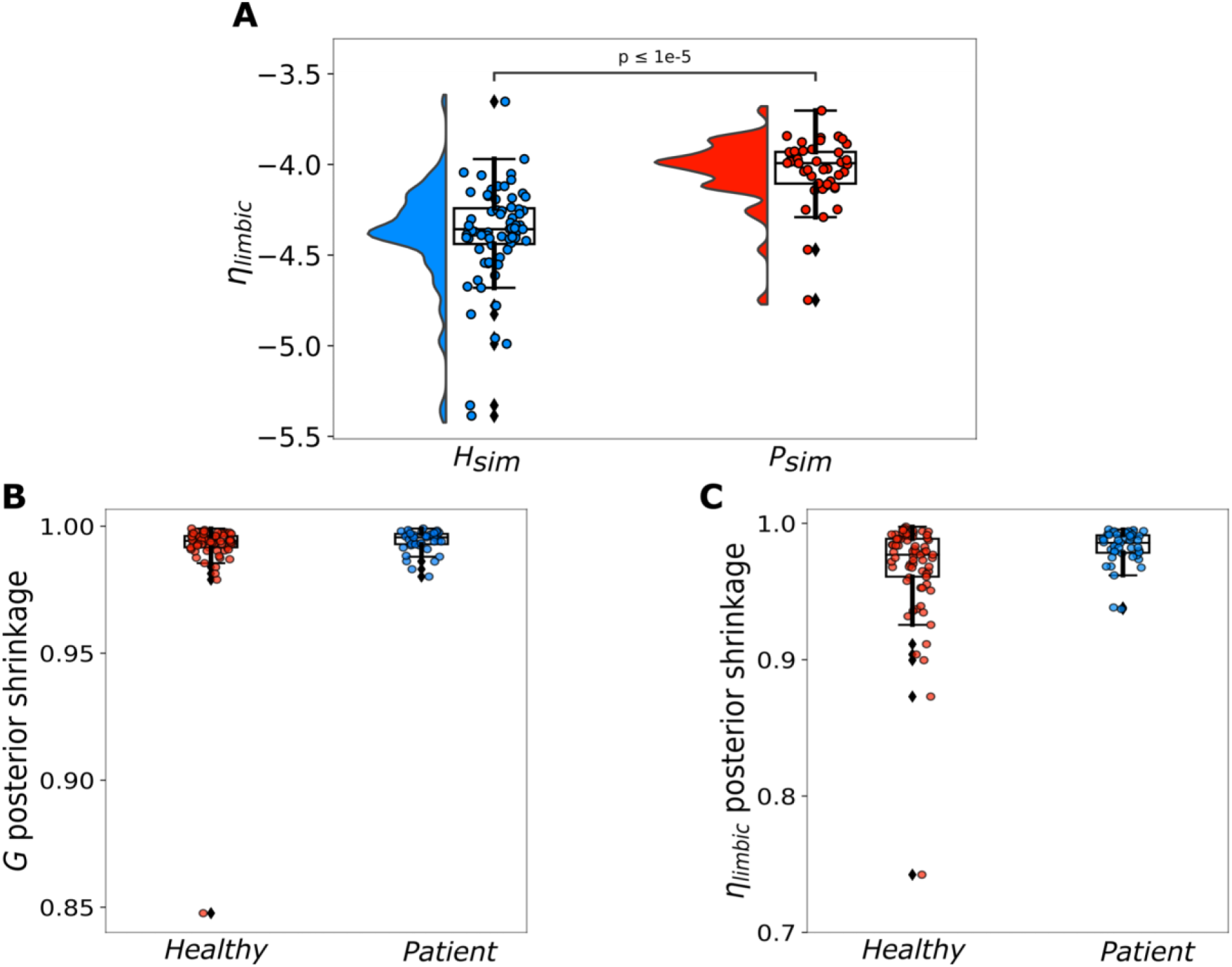
Simulation-based inference for parameters of virtual brains in healthy and AD groups. Global coupling and excitability in the limbic regions are inferred in order to achieve the best fit with the summary statistics containing the homotopic connectivity and limbic fluidity. **A**. Excitability of limbic regions ***η*** significantly increased in patients compared to the control group (*p* ≤ *1e*^−*5*^). **B-C**. Posterior shrinkage for each subject as defined by*s* = *1* − ***σ***^*2*^ _*post*_/***σ***^*2*^_*prior*_, where *σ*^*2*^_*prior*_ and *σ*^*2*^_*post*_ indicate the variance (uncertainty) of the prior and posterior distributions, respectively. The concentration of posterior shrinkages towards one indicates that all the posteriors in the Bayesian inference are well-identified, whereas its concentration towards zero indicates the poorly identified values.

The estimated joint posterior distribution using SBI for 9 randomly selected subjects (H_1-3_ representing the healthy subjects, P_1-3_ representing subjects from aMCI, and P_4-6_ representing subjects from AD group) are shown in Supplementary Figure S8. Moreover, the posterior shrinkage for each subject defined by *s* = *1* − *σ*^*2*^ _*post*_/*σ*^*2*^_*prior*_, are used as a diagnostic for SBI validation against empirical data. Here, *σ*^*2*^_*prior*_ and *σ*^*2*^_*post*_ indicate the variance (uncertainty) of the prior and posterior distributions, respectively (Figures 4B, C). The posterior shrinkage quantifies how much the posterior distribution contracts from the initial prior distribution. Hence, the concentration of estimated posterior shrinkages towards one indicates that all the posteriors in the Bayesian model inversion are well-identified.

## Discussion

In this study we employed computational brain modeling as a means of establishing a BNMs to explain abnormal brain activity observed in individuals with pathology, AD. To do this, we created a personalized BNMs in which structural brain topographies of each individual combined with disease related mechanisms, excitotoxicity and postsynaptic depression which are thought to be leading mechanisms of AD on neural activity. The mechanisms are applied in various levels considering one of the locations that has the highest protein burden, limbic network (Figure 1A). We discovered that excitotoxicity plays a critical role in reproducing the functional connectivity patterns seen in AD, particularly the hyperexcitation and impaired oscillations that may be caused by protein burden. Our findings showed that only hyperexcitation is sufficient and necessary to reproduce the empirical biomarkers of AD pathology, such as homotopic dysconnectivity and decreased limbic network dynamical fluidity which was not only observed in the diseased group, but also in healthy subjects who were virtually induced with excitotoxicity (Figure 3E, F). These results suggest that excitotoxicity may be a key contributor to the development of AD pathology and may be a potential therapeutic target for addressing this condition. Additionally, we demonstrated that our BNMs can estimate the relationship between excitability of limbic subnetwork and function using the Bayesian framework and a low-dimensional set of empirical summary statistics. We estimated the pooled posterior distribution of excitability parameter of limbic regions for each subject where the overall mean trend of *η* showed an increase with pathology (Figure 4A). Additionally, pooled estimated posterior distributions of parameters *G* showed non-significant change (Supplementary Figure S6). This was expected since no structural alterations were observed in patients’ connectomes rather than a decrease in the limbic network connections (Figure 2A, Supplementary Figure S1 and Supplementary Figure S2). Overall, our results show that the excitability of limbic local population activity in limbic network is the fundamental way to capture the properties of AD empirical data. By combining neuropsychological BNMs with simulation-based inference, we are able to provide a mechanistic explanation for the functional trajectory of AD.

These findings were obtained from analyzing data from 115 patients, including 71 healthy controls, 32 individuals with aMCI, and 12 individuals with AD, all of whom were between 70 and 90 years old.

Specifically, we introduced the regional heterogeneity to whole-brain dynamics through tuning the regional excitability of local population in the limbic subnetwork. By doing this, we were able to reveal the causal role that the limbic population plays in whole-brain neuronal dynamics (as measured by homotopic FC) and its relative impact over time in the limbic population (as measured by the variance in FCD as a metric for brain fluidity). To do this, we used a neural mass model that was inverted by fitting the parameters responsible for global modulation parameter and excitability in limbic regions. By integrating personalized information in BNMs, we are able to tune each model to maximize the variance of whole-brain FCD, *σ*_*full*_ ^*2*^, as a hypothesized dynamical working point which is considered representative of realistic dynamics of real data (Hansen et al., 2015; Rabuffo et al., 2021, Lavanga et al, 2022). Since global modulation is a measure of the influence of overall input mediated by SC, it reflects the relationship between structural and functional connectivity, or how changes in structural connections between brain regions (as measured by SC) can affect their functional interactions (as measured by FC). By manipulating the global modulation, it is possible to examine the effects of changes in SC on FC and brain fluidity. Unlike in healthy aging where this increases to effectively compensate for the lost SC (Lavanga et al., 2022, Petkoski et al, 2022), here there is no difference between the optimal G for both groups. This is not surprising given that SC is not different between the groups, Figure S1, and is additionally confirmed with the model inference, Figure S7. On the other hand, we demonstrate that the dynamics of an excitatory network can be altered by changing the heterogeneous inputs to the model, specifically the average excitability parameter of the mass neurons. By doing this, we are able to reproduce the empirical properties of static FC and FCD. Specifically, we find that increasing the average excitability in the limbic subnetwork of the mass neurons supports the hypothesis of hyperexcitation in the brain, which may be caused by the excitotoxic effects of pathological proteins such as Aβ and pTau in cortical regions. While our study does not have access to in-vivo quantitative PET imaging data that would allow us to map heterogeneity at the regional level, we were able to mimic hyperexcitability in the brain using brain network models by tuning the average excitability of the mass neurons in the limbic subnetwork. This choice of limbic subnetwork is based on the knowledge that pathological proteins such as Aβ and pTau are associated with hyperexcitability at different stages of the disease on those areas, and may even have synergistic interactions during disease progression (Roberson et al. 2007, Ittner et al. 2010, DeVos et al. 2013, Huijbers et al. 2019, Das et al. 2018, DeVos et al. 2018, and Sohn et al. 2019). Previous research has shown that individual with AD and MCI tend to have decreased functional connectivity in the default mode network in individuals with AD and MCI (Petrella et al. 2011, Agosta et al. 2012, Binnewijzend et al. 2012, and Pievani et al. 2014). However, more recent resting state studies demonstrated that these individuals also have decreased functional connectivity at the regional level in the limbic structures (Canuet et al. 2015) and at the whole-network level (Badhwar 2017). Our analysis suggests that the recorded empirical pathology-related homotopic dysconnectivity and reduced brain fluidity to flow between different cognitive states, which are important for cognition, are both driven by local hyperexcitation.

In individuals with AD, SC and FC is often disrupted, which is associated with clinical manifestation of disease. Graph analysis and connectomics are methods used to evaluate global and local, structural and functional properties of topological networks, as well as regional connectivity. These methods involve using neuroimaging techniques, such as MRI, EEG and MEG to study the connections between different brain regions in order to understand how they function and interact with one another. The connectomics analysis helped to understand the link between pathological changes and clinical symptoms in AD, and these connectome-based researches provide useful insight into progression of the disease. Pathologically, two main aberrant structures, Aβ and pTau, precede abnormal functional activity that shows cognitive impairment, memory loss, and loss of executive functions. Aβ progressively appear starts in the neocortex and continues through deeper brain areas during the development and progression of the disease, on the other hand pTau that also propagate, starts at the transentorhinal region and expands through the entorhinal cortex and limbic areas and eventually neocortical areas (Braak and Braak, 1991). Topographical changes generally computed from white-matter tractography or other methods while aberrant accumulated structures PET imaging of amyloid or biochemical analysis of the CSF. To assess functional integrity of communication between two regions, rs-fMRI is a great tool to reveal co-varying activity. FC can be constructed from pairwise Pearson correlation of the average fMRI (BOLD signal) time series for each region. Abnormal FC is detected generally in protein burden areas particularly in brain networks such as the default mode network, limbic network and the executive control network areas. Static FC analysis from rsfMRI is highly consistent, but FCD may provides a clearer working point for fitting virtual models (Battaglia et al. 2020; Lavanga et al. 2022, Petkoski et al., 2022) because it captures the non-stationary nature of brain activity. There is strong evidence that incorporating oscillatory dynamical properties in addition to static properties can improve the model optimization process. Researchers have identified a reduction in the “dynamic repertoire” of brain activity in AD, as indicated by disruptions in the evolution of FCD such as significant variations in dwell-time (Jones et al. 2012), a reduction in the functional states of global metastability (Cordova-Palomera et al. 2017), and a progressive loss of whole-brain metastability (Demirtas et al. 2017).

In our study, we found lower levels of FCD variance in the limbic areas (Figure 2C) as well as inter-hemispheric homotopic functional dysconnectivity (Figure 2B) in empirical data causally explain the data features observed in cognitively impaired with pathology groups. Both in healthy aging and AD, the functional homotopic disconnectivity has been related to corpus callosum degeneration in terms of volume reductions and diffusivity disruptions (Zhou et al., 2021; Zhang et al., 2021). The corpus callosum connects homologous regions of the cortex, and its degeneration may contribute to dysconnectivity in the transfer of information through inter-hemispheric homotopic connections, leading to memory impairment. Reduced homotopic functional connectivity is thought to be a key feature of AD, particularly in the temporal-parietal regions. Additional graph theory studies have indicated that the largest homotopic module and the insula module in individuals with AD have lost symmetric functional connectivity (Chen et al. 2013). Although the degeneration of corpus callosum, a major commissure of homologous connections between hemispheres, correlates with homotopic inter-hemispheric functional dysconnectivity (Qiu et al., 2016), there is no direct evidence that suggests homotopic disconnection is topographically driven. To determine whether recorded reductions in homotopic and FCD variance may be driven by network-based atrophy that disrupts large-scale network interactions in terms of functional specialization and segregation, we used a mask approach to replicate disease-related dynamical responses in silico. In this approach, we focused on network-based structural neurodegeneration, specifically the within limbic network and postsynaptic connections, which has been shown to have significantly reduced fractional anisotropy (FA) and increased mean diffusivity values (Pievani et al. 2010, Huang et al. 2012, Acosta-Cabronero et al. 2012, and Douaud et al. 2013). The structural damage in these regions has been found to correlate with the severity of symptoms and disease progression in AD (Pievani et al. 2010, Huang et al. 2012, Acosta-Cabronero et al. 2012, and Douaud et al. 2013). The mask approach incorporated empirical evidence of within limbic network atrophy driven by white matter deterioration and partially by the theory of tau epidemic, which suggests connectivity-mediated tau spreading (Vogel et al. 2020 and Frontzkowski et al. 2022). We selected the spread of tau (postsynaptic) in communicating neurons because white matter atrophy is commonly observed in limbic regions, which are particularly vulnerable to pTau accumulation and spread. There is strong evidence that increasing Braak and Braak pTau stages are correlated with increasing severity of white matter hyperintensities (WHM) (Erten et al. 2013). While the precise biological mechanism is not yet fully understood, WHM causes demyelination and axonal loss, leading to severe WHM damage in AD (McAleese et al. 2015). Therefore, testing the tau-spreading hypothesis through the limbic regions and postsynaptic connections using the mask approach can provide insights into the complex spatiotemporal dynamics of this process. We used this pathological-anatomical driver to reproduce the disease trajectory for each patient’s brain by masking the limbic network within and postsynaptic connections (inspired from Lavanga et al., 2022). Our brain network models verified that structural connectivity changes alone were not sufficient to exhibit the brain’s functional reorganization during disease, suggesting that the structure-function dichotomy does not display the same declining trends as the empirical data (Figure 3C, D).

Brains with pathology that reflect pathological functional reorganization virtually replicate empirical findings, later confirmed by deep learning networks used in the SBI approach (tailored to Bayesian framework). To estimate the parameters of BNMs, the optimization methods (within the Frequentist approach) have been often used in previous studies, by defining an objective function to score the performance of the model against the observed data. However, such a parametric approach results in only a point estimation, and the optimization algorithms may easily get stuck in a local minimum, thus, requiring multi-start strategies to address the potential multi-modalities. Using such classical methods, the estimation depends critically on the form of the objective function defined for the optimization process. Moreover, incorporating prior information (such as anatomical or functional data) into optimization problems can be challenging because it requires carefully balancing the trade-off between the fit to the data and the fit to the prior information. These issues can be solved by using the Bayesian framework. For instance, it provides a principled method to incorporate the prior knowledge through the use of a probability distribution over the model parameters to maximize the model predictive power. However, Bayesian estimation on high-dimensional and complex models such as BNMs with the large amount of data that is typically available for brain imaging studies is challenging. Particularly, in the presence of metastability in the state-space representation, Monte Carlo methods require either more computational cost or intricately designed sampling strategies to accurately detect state transitions in the latent space. To sidestep these issues, we used SBI for flexible and efficient Bayesian model inversion that readily estimate the full posterior of brain parameters from a low-dimensional set of empirical summary statistics (Cranmer et al., 2020; Gonçalves et al., 2020; Hashemi et al., 2020, Lavanga et al., 2022). These data features for accurate posterior distribution of global coupling and eta parameters included the homotopic dysconnectivity and limbic network fluidity decline, extracted from a limited number of BNMs simulations. We first demonstrated that SBI is able to recover the ground truth parameters, the global modulation, and excitability parameter for limbic subnetwork areas from whole-brain activities. Then by using this approach on empirical data, we found that the limbic areas excitability parameter significantly increases in AD patients relative to the control group, whereas significant changes were no longer occurring for the global modulation scaling the impact of subject-specific connectome.

## Conclusion

In conclusion, our research has provided insight into the connection between abnormal brain activity and the crucial role of the hyperexcitability i.e. excessive and hypersynchronous neuronal firing. We have found a causal link between regional variability of excitatory dysfunction which is mainly caused by protein burden, and global brain dynamics. Our results support the theory that protein pathology precedes structural changes and showed relation to functional network abnormalities and cognitive dysfunction. These key results emphasize the significant role that hyperexcitability plays in contributing to abnormal brain activity patterns in AD. On the other hand, postsynaptic depression, within the limbic subnetwork and postsynaptic connections, cannot be responsible for the observed functional differences. Bayesian approach using simulation-based inference, allows us to make efficient predictions about the behavior of individual brains by simulating them and comparing the results to the empirical recordings. In our research, we used deep neural density estimators to validate the change in limbic subnetwork regions excitability in AD. By estimating the full range of parameters, including global modulation and excitability in the limbic subnetwork, we were able to independently verify the change in excitability. Further research is needed to fully understand the relationship between functional network abnormalities and topography, and to explore the possibility that white matter degeneration may contribute to abnormal brain function. However, our work has contributed important knowledge to the field and has emphasized the importance of studying protein burden in the context of AD.

## Material and Method

a. Participants
b. Image Data Acquisition
c. Data Preprocessing
d. Structural, Functional Connectivity analysis and functional connectivity dynamics
e. The brain network model
f. Surrogate/Virtual models
g. Simulation-based inference

### M.1 Participants

The subjects in this study were drawn from the Sydney Memory and Aging Study (MAS) (Sachdev et al., 2010; Tsang et al., 2013), which included 115 community-dwelling participants aged 70-90 years. These subjects were stratified into three groups based on cognitive performance: 71 healthy controls (HC), 32 with amnestic mild cognitive impairment (aMCI), and 12 with Alzheimer’s disease (AD). The subjects underwent comprehensive neuropsychological assessments, as well as the Mini-Mental State Exam (MMSE) and the National Adult Reading Test (NART IQ) to screen for dementia and estimate premorbid intelligence levels, respectively. The neuropsychological assessments included tests of attention/processing speed, memory, language, visuospatial ability, and executive function (for a detailed description of neuropsychological tests, see, Zimmerman et al., 2018).

### M.2 Image Data Acquisition

Magnetic resonance imaging (MRI) scans, including structural diffusion (MRI) and resting-state functional MRI (rs-fMRI), were conducted using a Philips 3T Achieva Quasar Dual scanner. A single-shot echo-planar imaging (EPI) sequence was used for diffusion MRI, with a TR of 13,586 ms, TE of 79 ms, 61 gradient directions (b=2400 s/mm2), a non-diffusion-weighted acquisition (b=0 s/mm2), a 96×96 matrix, and a field of view (FOV) of 240×240mm2. Slice thickness was 2.5mm, yielding 2.5mm isotropic voxels. For rs-fMRI, a T2*-weighted EPI sequence was used with a TR of 2000 ms, TE of 30 ms, flip angle of 90°, FOV of 250mm, and a 136×136mm matrix size in Fourier space. 208 volumes were acquired, each consisting of 29 4.5mm axial slices. For structural connectivity analyses, a T1-weighted image was acquired with a TR of 6.39 ms, TE of 2.9 ms, flip angle of 8°, matrix size of 256×256, FOV of 256×256×190, and 1×1×1mm isotropic voxel slice thickness.

### M.3 Data Preprocessing

Diffusion magnetic resonance imaging (dMRI) preprocessing and whole-brain tractography were conducted using the same methods that were applied to a subset of the healthy control population (Perry et al., 2015; Roberts et al., 2017). Head motion correction was performed by rotating the gradient directions (Leemans and Jones, 2009; Raffelt et al., 2012), followed by bias field correction to reduce spatial intensity inhomogeneities(Sled et al., 1998). Fiber orientation estimation and whole-brain tractography were performed using the Mrtrix software package (v0.3.12-515; https://github.com/MRtrix3) (Tournier et al., 2012)., with constrained spherical deconvolution (CSD) (lmax=8) (Tournier et al., 2008) of diffusion-weighted MRI used to extract the fiber orientation density function (FOD). The probabilistic second-order integration over fiber orientation distributions (iFOD2) algorithm was used to propagate 5 million fiber tracks from random seeds, with a step size of 1.25mm, minimum track length of 12.5mm, maximum length of 250mm, FOD threshold of 0.1, and curvature of 1mm radius.

For fMRI data processing SPM8 function is used via Data Processing Assistant for Resting-State fMRI (DPARSF, v 3.2) (Chao-Gan and Yu-Feng, 2010). The following preprocessing steps are applied Slice-timing correction (realignment to mean functional image), co-registration to the structural image (via 6 DOF), linear detrending, nuisance regression of 24 motion parameters (Friston et al., 1996), and segmented WM/CSF signals (Ashburner and Friston, 2005). Generated average structural brain templates across all participants and native functional images were transferred to MNI space (3 mm) via this template. Lastly, smoothing (8 mm) and temporal band-pass filtering (0.01–0.08 Hz) was employed.

For functional magnetic resonance imaging (fMRI) data processing, the Data Processing Assistant for Resting-State fMRI (DPARSF, v 3.2) software package was used in combination with SPM8 (Chao-Gan and Yu-Feng, 2010). The fMRI data underwent several preprocessing steps, including slice-timing correction (realignment to mean functional image), co-registration to a structural image, linear detrending, nuisance regression of motion parameters, segmentation of white matter and cerebrospinal fluid signals, transformation to the Montreal Neurological Institute (MNI) space, and smoothing and temporal band-pass filtering. Pre-processing pipeline of these data described in detail elsewhere (Perry et al., 2017).

### M.4 Structural, Functional Connectivity and Functional connectivity dynamics Analysis

Interareal connectivity estimates (Tzourio-Mazoyer et al., 2002) of SC and FC were built via parcellating the whole-brain into widely-used AAL Atlas (within MNI space) comprising 6 lobes; frontal, temporal, parietal, occipital, insula-cingulate and central structures (Wang et al., 2006; Savio and Graña, 2017; Tijms et al., 2018). Subject-space parcellation was achieved using FSL5. Individual FA images were linearly co-registered into the FMRIB standard space, and the AAL parcellation was subsequently transformed into subject-space by applying the inverse of this transformation matrix. Quality control procedures for SC are described in detail elsewhere (Zimmerman et al., 2018). For SC matrices, a connection matrix (*W*_*ij*_) representing the total number of streamlines (with a 2mm radius) between regions i and j, and a distance matrix were assessed using tractography. The 2mm radius was selected as the default parameter for MRtrix’s tck2connectome function, which also identified erroneous fiber terminations as connectome edges. This streamline identification method prevents potential false negatives and under-sampling of connectome edges. The total number of streamlines was corrected using the euclidean distance and the total volumetric size (summed number of voxels) of the two regions. As described in Zimmerman et al., 2018, a subset of the AAL regions was used for the limbic network model.

To construct the FC matrices, we calculated the pairwise Pearson correlation of the average fMRI (BOLD signal) time series for each region (i.e., all region i voxels between all ij region pair). This resulted in a symmetric matrix with 88 nodes, where each entry represents the Pearson correlation coefficient between the two respective nodes.

In particular, we selected biomarkers that reflect characteristic changes in the intrinsic functional architecture in AD and aMCI. The average homotopic FC strength is the average sum of the connections between the two hemispheres:

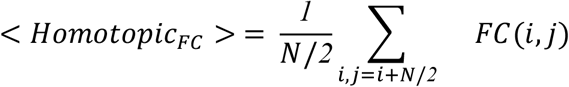

where the averaged *FC*(*i, j*) entries represent the element of order 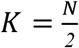 (Mollink et al., 2019). To further investigate the dynamical changes we computed the time-variant representation of FCD using a sliding window Pearson correlation coefficient (Battaglia et al., 2020) which reflects a capacity of brain’s flexibility (Córdova-Palomera et al., 2017).

The window size was set to 40 seconds with maximum overlap, which is sufficiently sensitive to demonstrate the temporal transitions of the FC streams (Jia et al., 2017; Lurie et al., 2020). The similarity of any two FC is represented via a time-time matrix, in which *FC*_*t1*_ and *FC*_*t2*_ are introduced as upper triangle Pearson correlation between entries:

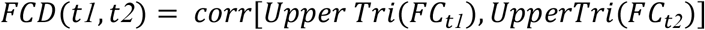

We quantified the switching behavior of the resting state networks (also known as the switching index, SI) by calculating the variance of the upper triangular part of the respective FCD matrix (Hansen et al., 2015; Rabuffo et al., 2021):

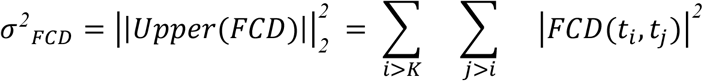

where upper triangle matrix diagonal order is *k* = *widow size* to correct for overlap. The SI reflects the degree to which the neural activity of nodes switches between “up” and “down” states.

### M.5 The brain network model

We simulated resting-state activity using the neural mass model (NMMs) introduced by Montbrió et al., 2015. This model represents the dynamics of a brain region as an ensemble of infinite, all-to-all coupled quadratic-integrate-and-fire (QIF) neurons at the thermodynamic limit. The model assumes that the dynamical currents are distributed according to a Lorentzian distribution, resulting in a system of firing-rate equations composed of two ordinary differential equations:

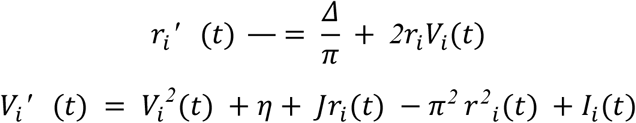

where *r*_*j*_ (*t*) is the mean population firing rate at time t, *V*_*i*_(*t*) is the mean membrane potential, *J* is the average synaptic weight, *η* is the average excitability of the mass neurons, and Δ is the spread of heterogeneous noise. This model has been derived as the exact limit of an infinite number of all-to-all coupled quadratic integrate-and-fire (QIF) neurons according to the Ott–Antonsen ansatz. The firing rate attempts to characterize the action potentials generated by a population of spiking neurons in an excitatory network, with both heterogeneous inputs *η* and the synaptic weights *J*. The parameter default values were set to *J* = *14*.*5*, Δ = 0.7 and η = −4.6 to place each decoupled node in its bistable regime, where the 2D phase space demonstrates a ‘down’ fixed point and an ‘up’ stable focus (Rabuffo et al., 2021). This realistically represents the switching behavior observed in empirical data (Hansen et al., 2015; Rabuffo et al., 2021).

The impact of the SC over the global dynamics is then scaled by the parameter G in the additive current input *I*_*i*_(*t*) :

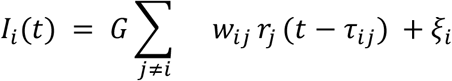

where weights of the SC edges are represented by *w*_*ij*_, and the stochastic noise by ξ_*i*_ that allows the region to oscillate between the up→down or down→up states. We used the Virtual Brain platform with the Heun-stochastic integration to simulate the activity of each brain region. The variables *ri* (*t*), and *V*_*i*_(*t*) of each region were downsampled by sampling frequency *fs* = *100 Hz*. The simulated BOLD activity for each node was calculated by filtering the membrane potential using the Windkessel model, which emulates fMRI time series with a repetition time of 2000 ms.

### M.6 Virtual brain model for AD

One of the hallmarks of AD is an abnormal increase in neural excitability, which is thought to contribute to the cognitive decline and neurodegeneration seen in the disease. This hyperexcitation can occur at the cellular level, where neurons become more likely to fire action potentials, and at the network level, where there is an increase in synchronous activity between neurons. This increase in excitability has been observed in various brain regions in AD, including the hippocampus, amygdala, and prefrontal cortex, and has been linked to the accumulation of beta-amyloid plaques and pTau tangles, which are characteristic features of AD. In this study, we introduce a novel approach to investigate the effect of local excitability on the ability of neural mass models to reflect dynamical changes. BNMs are often used to study the collective behavior of large populations of neurons, and changes in excitability can have significant impacts on the dynamics of these systems. By examining the influence of local excitability on the whole brain dynamics, we hope to gain a better understanding of the role of excitability in AD brain function.

The neural mass model (NMM) is a mathematical model that represents the collective behavior of large populations of neurons. It is often used to study the dynamics of brain activity and can provide insights into the underlying mechanisms of brain function. In this study, we used the all-to-all coupled quadratic-integrate-and-fire (QIF) neuron system, as described in Montbrió et al. (2015), to simulate the activity of each brain region. In this system, the excitability of each neuron is drawn from a Lorentzian distribution with density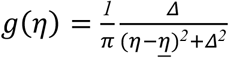. When the initial values are larger than the excitability threshold, the neuron exhibits spiking behavior. The exact dynamics of the system depend on the excitability and the size of the synaptic weight. (For details Montbrió et al., 2015).

In the virtual model neural mass model we fit the parameters global modulation index (*G* ∈ [*0*.*2, 1*.*1*]), noise variance *σ*^*2*^ ∈ [*0*.*01,0*.*05*] and excitability of limbic subnetwork regions *η* ∈ [−*6*.*0*, −*3*.*0*]). After noise parameter set to *σ*^*2*^ = *0*.*04*, we infer our connectomes for the most realistic switching index couple. Using this virtual model, we quantified the temporal evolution of simulated brain connectivity by comparing a set of static and dynamic FC properties to those captured in empirical data (as it’s described at M.4)

### M.7 Simulation-based inference

In order to retrieve the model parameter space compatible with the empirical data, the global modulation parameter *G* and excitability parameter *η* the limbic subnetwork was estimated using the Bayes-tailored approach known as simulated-based inference (SBI; Cranmer et al., 2020; Gonçalves et al., 2020). The Bayes theorem calculates the posterior distribution of parameters given observations *p*(*θ*|*y*), as a renormalized pointwise product of prior distribution *p*(*θ*), and the likelihood function of observations given model parameters *p*(*y*|*θ*), through: *p*(*θ*|*y*) ∝ *p*(*θ*) *p*(*y*|*θ*). The prior distribution *p*(*θ*) of a random event is unconditional probability of a data through beliefs before any evidence from observed data is taken into account, whereas the likelihood function *p*(*y*|*θ*) is the conditional probability of the observed data *y* given a certain set of parameter values *θ*. For many high-dimensional nonlinear models, such as the BNMs, the calculation of likelihood function becomes computationally expensive or prohibitive, hence, the likelihood approximation algorithms are required to efficiently estimate the parameters (Hashemi et al., 2022). SBI aims to perform flexible and efficient Bayesian inference for complex models when standard methodologies cannot be applied, due to analytic or computational difficulties in calculating the likelihood function (Cranmer et al., 2020). Sequential neural posterior estimation (SNPE; Gonçalves et al., 2020) is a class of SBI methods, which directly estimates the full high dimensional posterior density from forward simulations of a generative model (such as BNMs used in this work), bypassing the need for Monte Carlo sampling. SNPE requires three inputs (Gonçalves et al., 2020): (i) prior distribution describing the possible range of parameters from which the samples can be drawn easily, (ii) a mechanistic model that takes parameters as input and generates simulated data as output for training, and (iii) a set of observations (low-dimensional data features) as the target of fitting. By drawing parameters from prior *θ*_i_ ∼ *p*(*θ*), the simulated data set {(*θ*_i_, *y*_i_)}_i=1_ ^N^ was generated for training, where N is the number of independent and identically distributed samples from the generative model *p*(*θ, y*) = *p*(*θ*) *p*(*y*|*θ*), with *y*_i_ as the low-dimensional data features of forward simulations. By using a class of neural density estimators, such as masked autoregressive flow (MAF; Papamakarios et al., 2017) for training on the simulated set, then we are able to readily estimate the approximated posterior *qφ*(*θ*|*y*) with learnable parameters *ϕ*, so that for the observed data *y*_*obs*_: *qφ*(*θ*|*y*_*obs*_) ≃ *p*(*θ*|*y*_*obs*_). We ran a MAF in SNPE, with 5 autoregressive layers, each with two hidden layers of 50 units, as implemented in the PyTorch-based SBI package (Tejero-Cantero et al., 2020).

In this study, *θ*={*G, η*}, and *N* = *2*^*16*^ simulations were performed on the generative model, with uniform prior as *G* ∈ [*0*.*2, 1*.*1*] for the global coupling parameter, and *η* ∈ [−*6*.*0*, −*3*.*5*] for the excitability parameter of the limbic subnetwork. Then, the MAF was trained on a selected set of data features including the mean homotopic FC, and FCD variance within the limbic network. The final posterior estimation resulted in the joint probability distribution of *G* and *η*, given the data feature of empirical BOLD signals as *p*(*G, η* |*y*_*empirical*_). For SBI, the noise level was set to the optimized value obtained from the parameter sweeping.

### Data and materials availability

The Virtual Brain is publicly available (https://github.com/the-virtual-brain/tvb-root), while learning material to master can be found at the TVB hub website (https://hub.thevirtualbrain.org/). The Structural Connectivity data, fMRI time series and the cognitive scores are available upon reasonable request by sending an email to XX.

## Data Availability

All simulated data produced in the present study will be made available upon publication. 
All personalized data used in the present study are available upon reasonable request to the authors

## Acknowledgments

This project was funded by the European Union’s Horizon 2020 research and innovation programme under grant agreement No. 785907 (SGA2), and No. 945539 (SGA3) Human Brain Project, and Virtual-BrainCloud (grant number 826421).

## Supplementary materials

### SM-Connectome-based model with structural impact of protein burden on limbic network and postsynaptic links

To understand the association of white matter degeneracy, and protein accumulation on heterogeneous functional changes that distinguish the healthy brain from the patient’s brain we have designed different virtual brains. We test whether limbic network structural degeneration drives functional changes during progression healthy to clinically diagnosed as cognitively declined. First we extract patients structural adjacency matrix of the MAS study subject structural phenotype (*SC*_*emp*_). To test SC changes is the enough to convey on dynamical changes we focused on two in silico summary statistics emerging from the virtually aged BNMs with (i) subject structural phenotype (*SC*_*emp*_) hypothesized no explicit impact of protein burden neither on links nor on local excitability and (ii) hypothesized impact of protein burden on limbic network and postsynaptic links (*SC*_*mask*_) (from 0% to 100% decrease) (Lavanga et al., 2022).

For hypothesis two we performed virtual structural degeneration on aMCI and AD subject connectome by homogeneously decreasing the within limbic network and postsynaptic connections via mask approach (Lavanga et al., 2022);

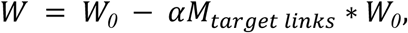

Where *M*_*target links*_ is the within limbic network and outgoing connection mask, *α* is the normalized intensity of decrease and * represents an element-wise product. In both only connectome based simulations and masking method the brain network model fitted by parameters *G* (*G* = [*1*.*5* − *3*.*2*], *with* Δ*G* = *0*.*05*) modulation index and noise variance *σ*^*2*^ (*σ*^*2*^ = [*0*.*01* − *0*.*05*] with Δ *σ*^*2*^ = *0*.*01*) to capture switching behavior of functional brain networks that reflects realistic evaluation of brain function. After having the most realistic switching index couple (*G, σ*^*2*^) for connectome based simulations and masking methods, the temporal evolution of simulated brain connectivity is quantified via a set of static and FCD properties that are captured in empirical data. Homotopic FC and limbic subnetwork FCD variance difference is used to check whether connectome-based simulated data, mask-simulated data could reproduce the dynamical and statical properties of empirical FC. As expected, BNMs in both approaches were not sufficient to capture the representative dynamics of each group (Figure 3A, B and Figure 3C, D). We also addressed a relationship between G modulation index and pathology. The structural connections between nodes influence the dynamic of each node and global parameter G represents the impact of structural architecture over the brain’s dynamics in the brain network model. Results showed that *G* modulation index was not associated with pathology, the whole-brain structural changes were not significantly different between healthy and patients brain. (Figure S3B,C)

**Figure S1.**
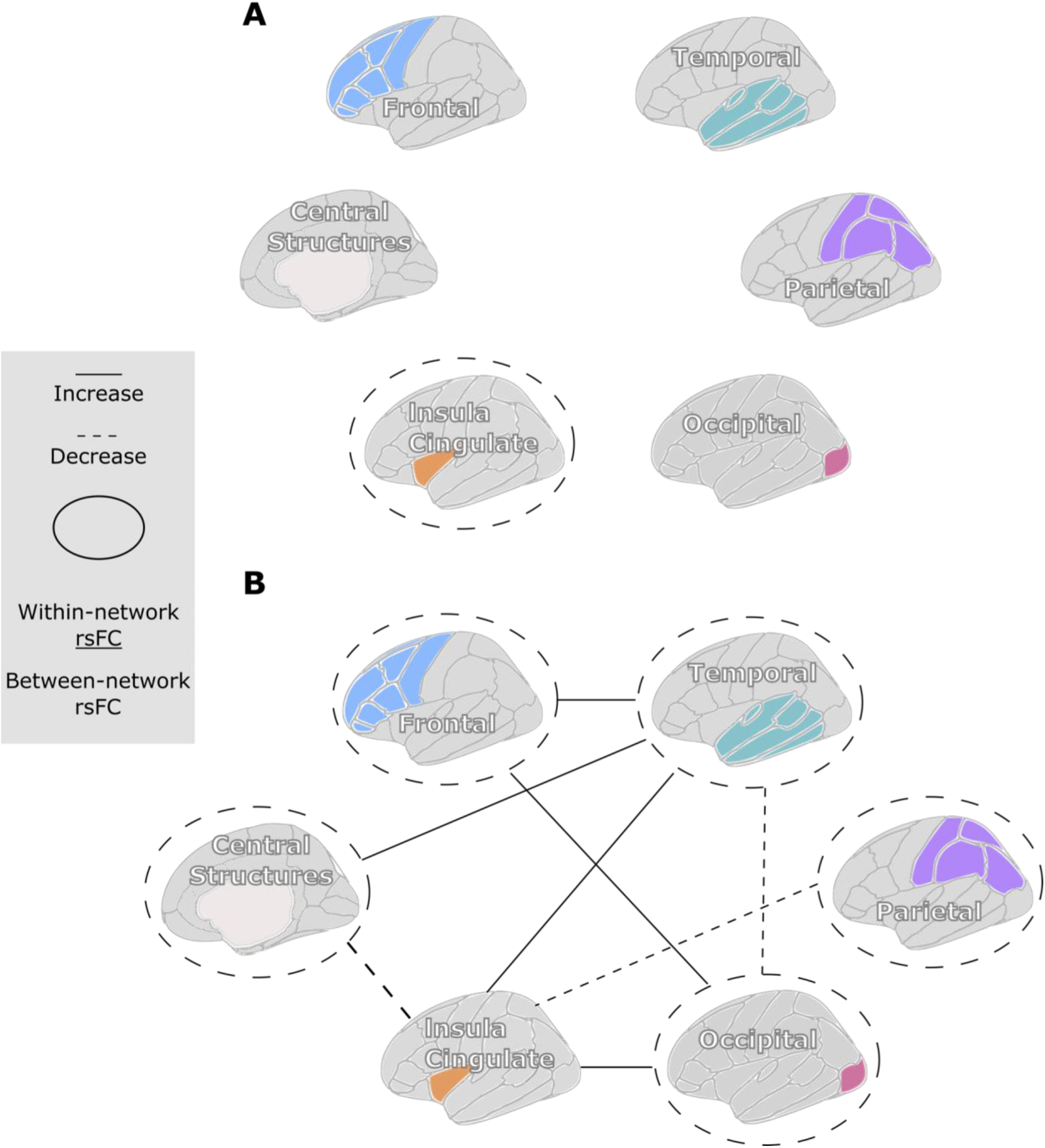
Within and between networks SC-FC. AAL atlas is divided into six distinct lobes: Frontal, temporal, parietal, occipital, insula cingulate and central structures. **A**. Only Insula Cingulate lobe connections showed decrease via disease which share regions with the limbic subnetwork. **B**. Shows functional reorganization due to disease. Within network connections significantly reduced in all lobes except the insula-cingulate lobe. Reductions between network connections were recorded for several connections: Central Structures-Insula Cingulate; Parietal-Insula Cingulate. The increase between network connections were recorded for several connections: Frontal-Temporal; Frontal-Occipital; Temporal-Insula Cingulate; Temporal-Central Structures; Insula Cingulate-Occipital.

**Figure S2.**
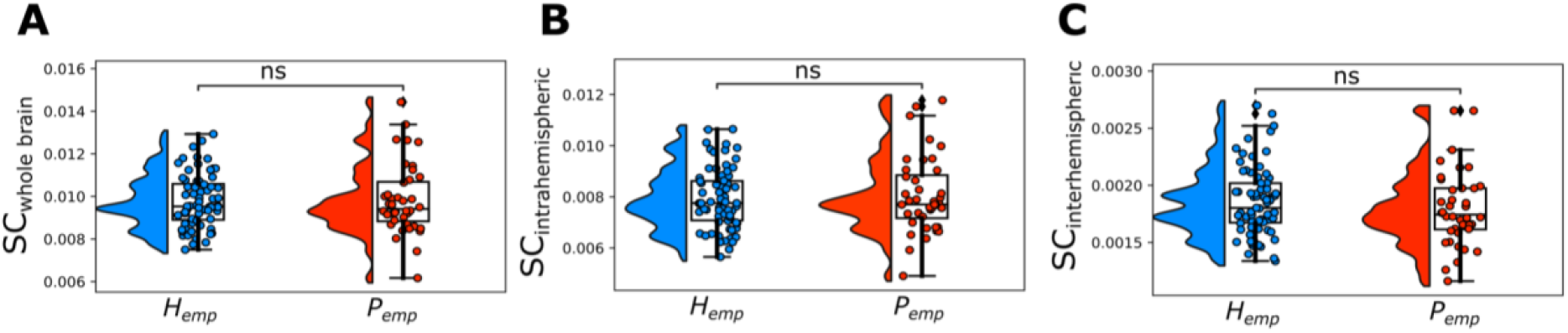
Comparison of structural connectivity data features between the groups. **A-C**, The average whole-brain, intra and inter hemispheric SC, respectively, for healthy control and cognitively impaired groups with pathology.

**Figure S3.**
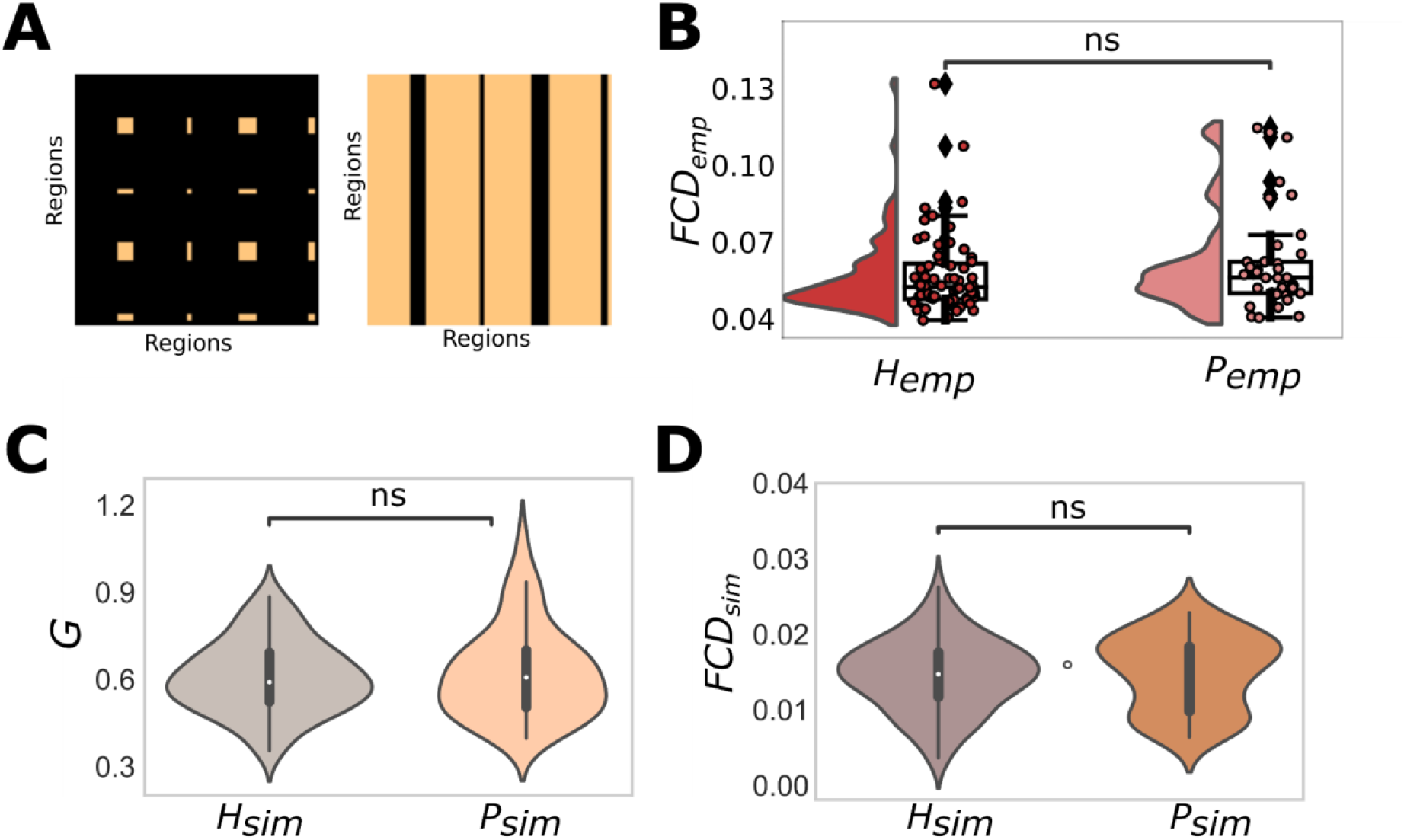
Limbic network within and polysynaptic connections mask and related whole-brain FCD and the global modulation G. **A**. Showing artificial decrease of within limbic structural SC by masking the within limbic network and postsynaptic connections of the SC matrix (from 0% to 100% decrease) for the testing SC-FC dichotomy in Alzheimer’s disease. **B**. The variance of whole-brain dynamics as measured by FCD shows no significant changes for empirical data. **C**. The global modulation associated with maximum FCD variance shows non-significant changes in the healthy control group compared to the cognitively declined group. **D**. The variance of whole-brain dynamics as measured by FCD also shows no significant changes. (**C-D** represents only connectome simulation)

**Figure S4.**
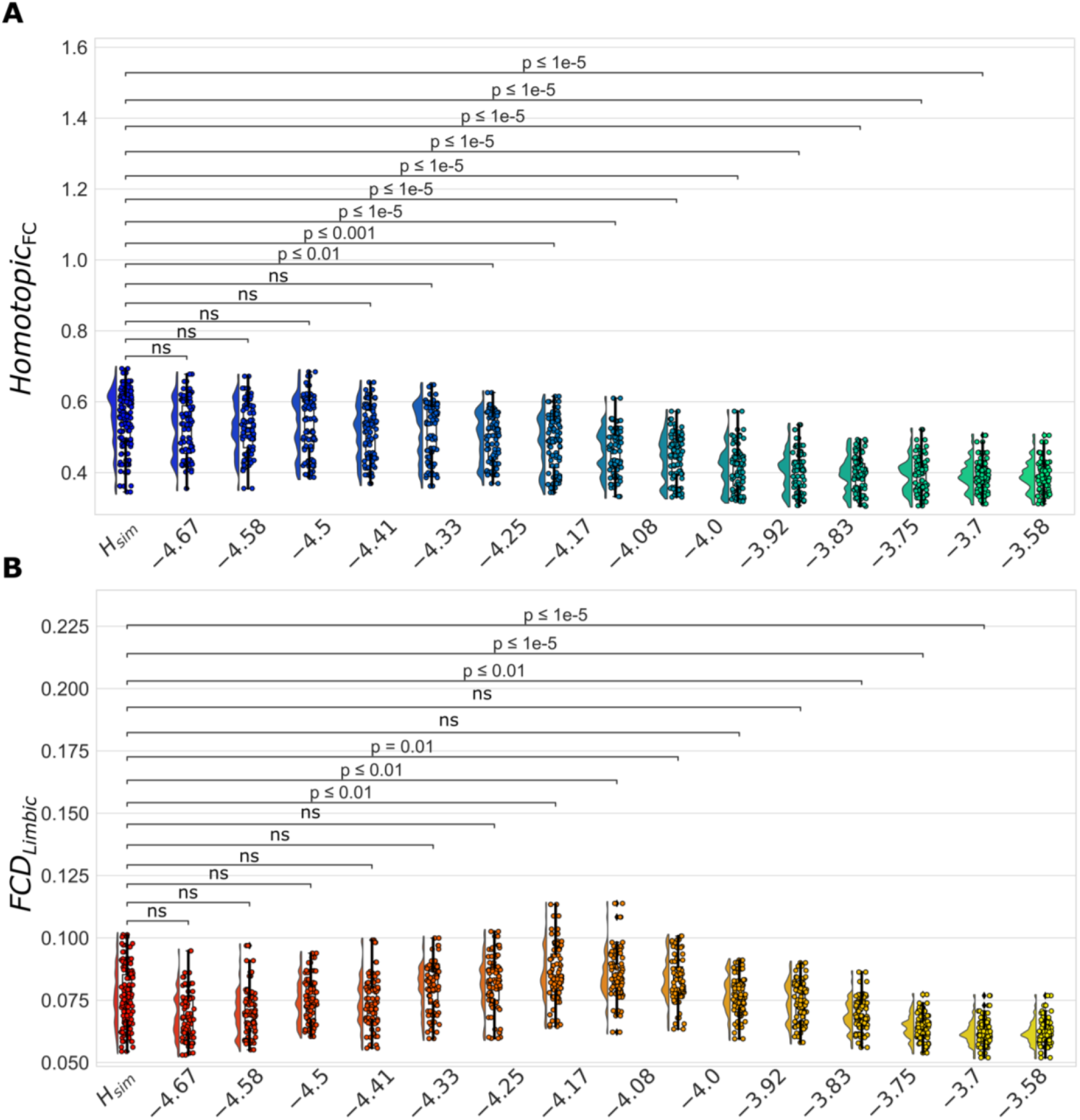
Simulations for varying 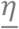 of limbic regions in MCI and Alzheimer’s subjects brain dynamics compared to healthy control with 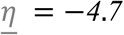. **A**. Homotopic functional connectivity decrease recorded as increased 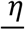 and **B**. reduced limbic network fluidity recorded after.

**Figure S5.**
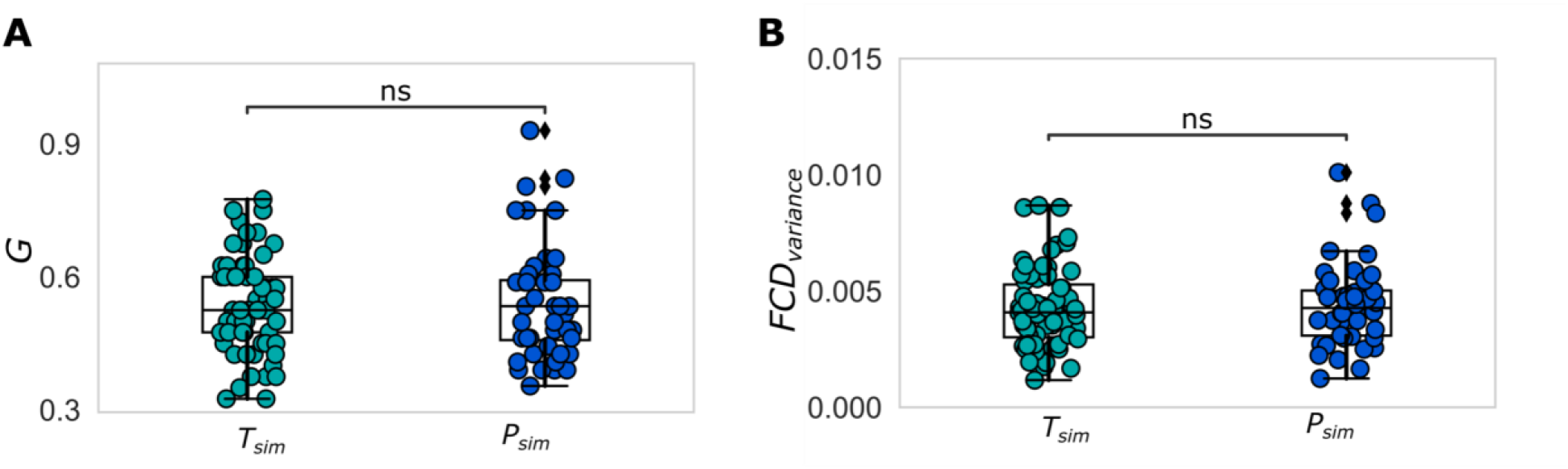
Global Modulation and Fluidity measures of whole brain dynamics for the working point of simulated brains with limbic excitability *η* = −*3*.*58*. **A**. The global modulation associated with maximum FCD variance shows non-significant changes in the patients group (*P*_*sim*_) and for the virtually diseased group consisting of the healthy subjects with hypothesized impact of the protein burden (*T*_*sim*_). **B**. The variance of whole-brain dynamics as measured by FCD also shows no significant changes patients group (*P*_*sim*_) and for the virtually diseased group consisting of the healthy subjects with hypothesized impact of the protein burden (*T*_*sim*_).

**Figure S6.**
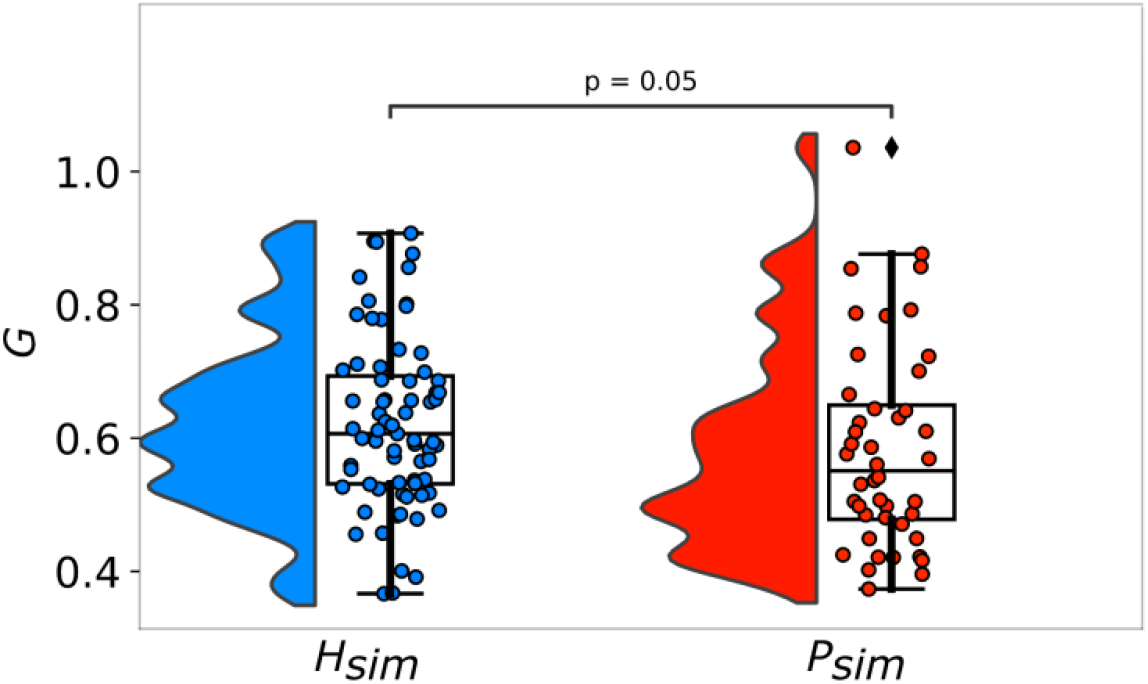
Pooled estimated posterior distributions of global modulation *G* shows non-significant change patients group (*P*_*sim*_) relative to the healthy group patients group (*H*_*sim*_) (*p* = *0*.*05*).

**Figure S7.**
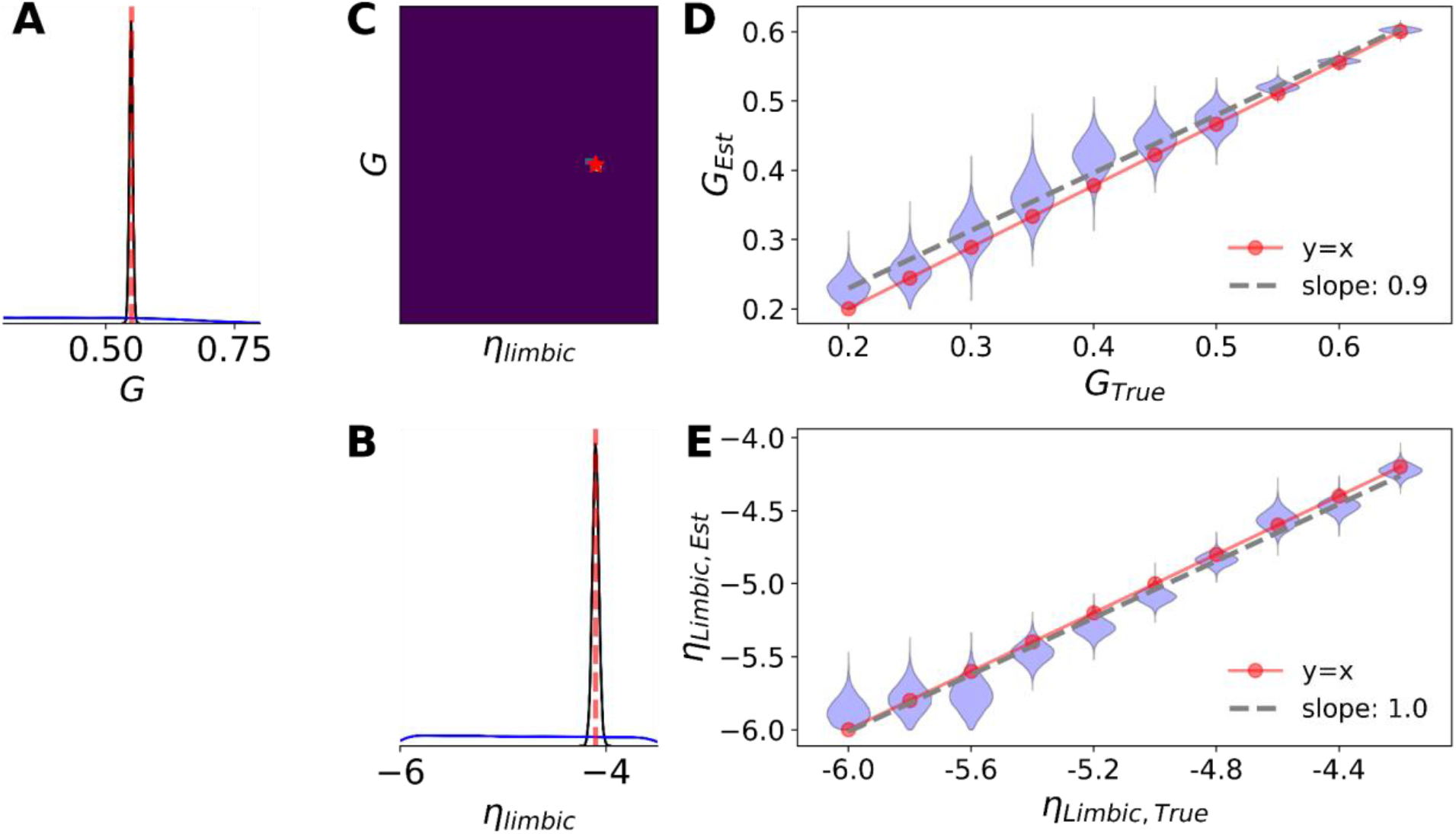
Validation of parameter estimation using a simulated BOLD time series as the observation, with known parameters. **A-B**. Inferred posterior distributions for the global coupling strength *G*, and the excitability of limbic subnetwork *η*, respectively, given low dimensional data features (summary statistics of FC/FCD). The ground truth parameters are shown in vertical red lines (*G* = *0*.*55* and *η* = −*4*.*1*). The neural network trained by 65k simulations with uniform prior (in blue) as *G* ∈ [*0*.*2, 1*.*1*] for the global coupling parameter, and *η* ∈ [−*6*.*0*, −*3*.*5*] for the excitability parameter of the limbic subnetwork, in a single round of SNPE. SBI accurately estimates a posterior distribution (in black) that narrows around the true parameters. **C**. Joint posterior distribution between parameters *G* and *η* (correlation=-0.7) demonstrates a very high posterior shrinkage, impling an ideal Bayesian inversion. **D-E**. As the brain dynamics is changed by varying the the parameters *G* and *η*, the inferred posterior distributions (violin plot, in blue) accurately encompass the true values (red circles). Dashed black lines represent a linear regression on mean estimated values, which demonstrate very close agreement with a perfect fit (y=x). Due to the amortization strategy (i.e., a single pass through the artificial neural networks, without the need for retraining), SBI is able to rapidly (in the order of a few seconds) estimate the posterior of parameters for new observations. These results validate the reliability and accuracy of SBI, for whole-brain model inversion from low-dimensional data features.

**Figure S8.**
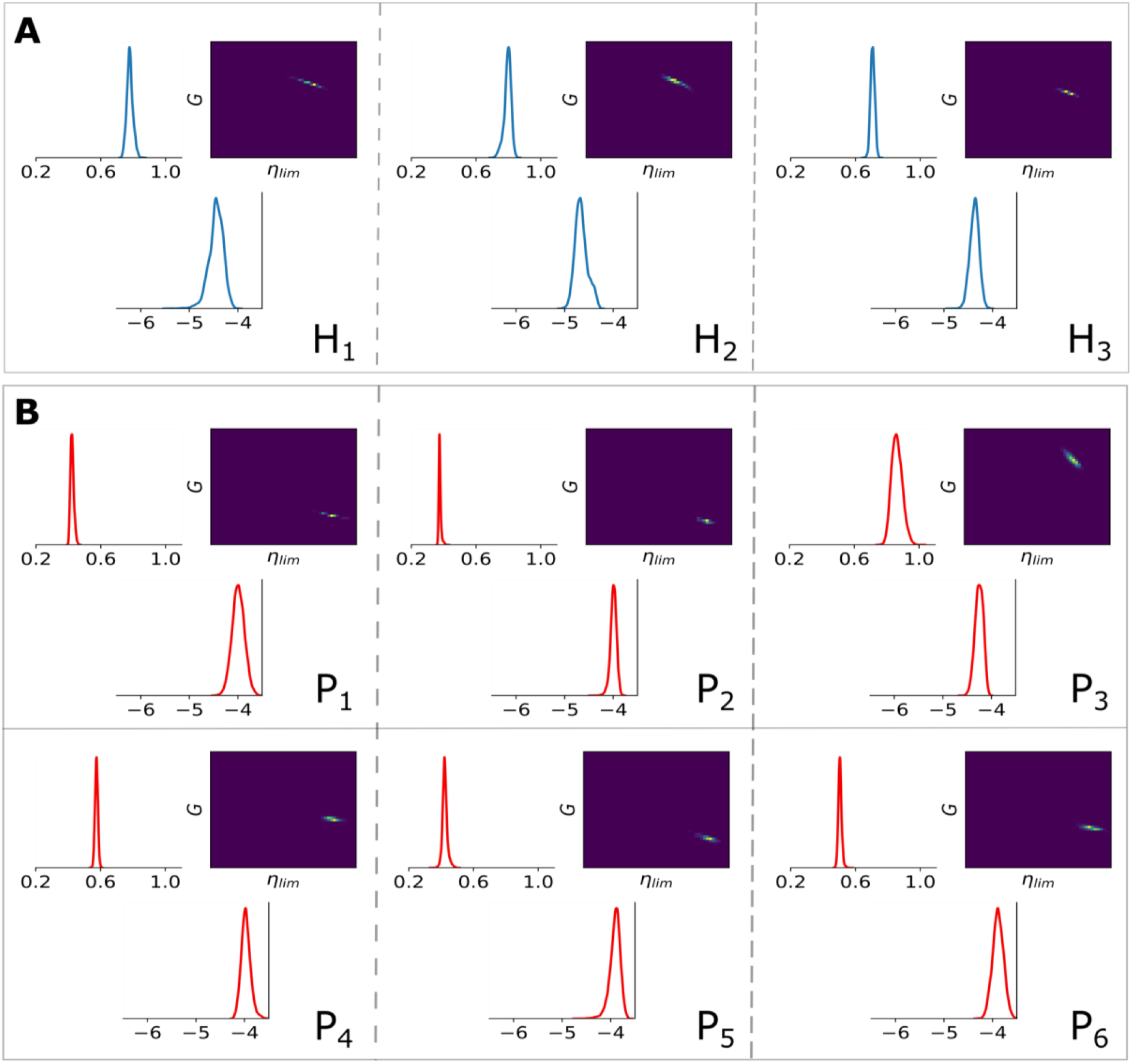
Joint posterior distribution of virtual brains in healthy and AD groups.Posterior estimation using SBI for 9 randomly selected subjects: **A**. H_1-3_ representing healthy subjects, **B**. P_1-3_ representing subjects from aMCI and P_4-6_ representing subjects from AD group. The neural network trained on ensemble of 65k simulations with uniform prior as *G* ∈ [*0*.*2, 1*.*1*] for the global coupling parameter, and *η* ∈ [−*6*.*0*, −*3*.*5*] for the excitability parameter, in a single round of SNPE.

**Figure S9.**
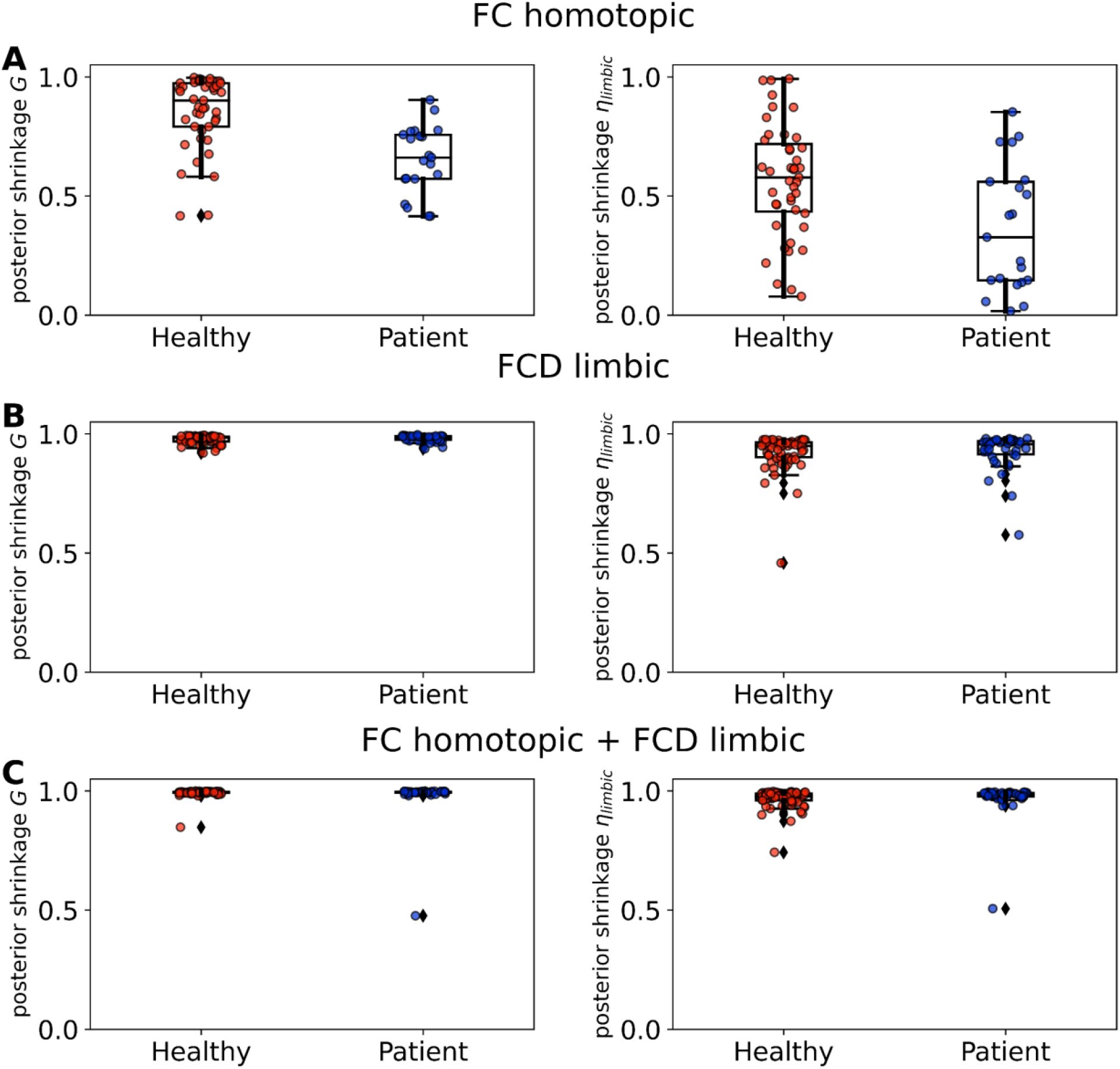
Comparing the posterior shrinkage of estimated global coupling *G*, and excitability *η*within limbic network, using summary statistics as: **A**. Only FC homotopic mean value of BOLD time series, **B**. Only FCD variance of the limbic subnetwork. **C**. both data features.

## Notes

### Competing Interest Statement

The authors have declared no competing interest.

### Author Declarations

Only existing public datasets were used. In particular it is data from Sydney Memory and Ageing Study. https://cheba.unsw.edu.au/research-projects/sydney-memory-and-ageing-study

